# Multi-transcriptomic analysis points to early organelle dysfunction in human astrocytes in Alzheimer’s disease

**DOI:** 10.1101/2021.02.25.21252422

**Authors:** Elena Galea, Laura D. Weinstock, Raquel Larramona-Arcas, Alyssa F. Pybus, Lydia Giménez-Llort, Carole Escartin, Levi B. Wood

**Author notes:** <u>R. Larramona</u>: Celltec-UB, Departament de Biologia Cel.lular, Fisiologia i Immunologia, Universitat de Barcelona, 08028; Institut de Neurociències, Universitat de Barcelona, 08035, Barcelona, Spain.

## Abstract

Recent single-nucleus RNA sequencing of astrocytes in *postmortem* Alzheimer’s disease (AD) samples is limited by the low number of sequenced astrocytes and small cohort sizes. We developed a systems biology approach to identify astrocytic genes in public large bulk AD transcriptomes. Brain cell specific gene clusters were generated from RNA sequencing data from isolated healthy human brain cells using cell-type enrichment scoring and clustering. Cell-specific gene clusters were localized in whole-brain transcriptomes from 766 subjects diagnosed with AD or mild cognitive impairment (MCI) from the Mount Sinai Hospital, the Mayo Clinic, and the Religious Order Study/Memory and Aging Project (ROSMAP). Gene clusters were organized into manually curated functional categories including astrocytic categories absent from existing platforms, such as perisynaptic astrocytic processes (PAP). Changes among subjects were determined by gene set variation analysis (GSVA) and principal component analysis (PCA). Hierarchical clustering revealed molecular heterogeneity in individuals with the same clinical diagnosis. Particularly in the Mayo Clinic and ROSMAP cohorts, over 50% of Controls presented the massive down-regulation of genes encoding synaptic proteins typical of AD, whereas 30% of AD patients presented Control-like transcriptomic profiles. According to GSVA and PCA, down-regulation of neuronal genes related to synaptic proteins correlated with down-regulation of astrocytic genes encoding mitochondrial and endolysosomal proteins, and up-regulation of genes related to PAP. Transcriptomic data thus unravel a deep phenotypic transformation of human astrocytes in AD, affecting organelles and astrocyte-neuron interactions. We posit that therapies preventing organelle dysfunction in astrocytes may protect neural circuits in preclinical and clinical AD.

**Main points:**

1. Transcriptomes of control subjects and AD patients segregate in two main molecular profiles regardless of clinical diagnosis.
2. Dysregulation of the endolysosomal/mitochondrial axis in astrocytes correlates with impaired synaptic plasticity in neurons.

## Introduction

‘Reactive’ GFAP-overexpressing astrocytes are found in the vicinity of amyloid-β plaques in *postmortem* brains from patients with Alzheimer’s disease (AD), in both the autosomal-dominant Alzheimer’s disease (ADAD) and the sporadic late-onset (AD) variants, as well as in mouse models of ADAD. The robust morphological transformation of astrocytes points to major phenotypical and, hence, functional alterations in astrocytes in AD (reviewed in (Perez-Nievas & Serrano-Pozo, 2018)).

However, despite non-negligible research in the last decade, knowledge about phenotypical alterations in human astrocytes from AD patients is still limited due, in part, to scarce and conflicting human astrocyte-specific omics data. Prior studies include transcriptomics of laser-microdissected astrocytes (Sekar et al., 2015; Simpson et al., 2011), co-expresion-based gene clustering of whole-brain AD transcriptomes (B. Zhang et al., 2013) (Neff, 2021), and single-cell RNA sequencing (scRNAseq) analyses (Grubman et al., 2019; Mathys et al., 2019).

Laser-microdissected astrocytes either presented insufficient number of differentially-expressed genes (DEG) for pathway analysis (Sekar et al., 2015), or normalization anomalies, as suggested by the finding that 98% of the DEGs were down-regulated (Simpson et al., 2011). The problem may lie in the low RNA yields of laser microdissection, exacerbated by the poor RNA quality after long *post-mortem* intervals. As an alternative to laser microdissection, clustering statistics based on gene co-expression identified function-specific gene modules using whole-brain AD transcriptomes (B. Zhang et al., 2013). Cellular localization of modules was established using cell-specific markers, and causality between nodes was then inferred with Bayesian statistics. Using this approach, alterations of glutamate and amino acid metabolism in an astrocytic module of 260 genes were discovered to be well-ranked for causal relevance in AD, but no further changes were unraveled. Finally, although scRNAseq and single-nucleus (sn)RNAseq improve the cellular resolution of transcriptomics to the point of unraveling populations within a given cell-type, two challenges arise in studying astrocytes with these techniques. One is the low numbers of astrocytes per patient that are sequenced, which renders astrocytic datasets underpowered (discussed in (Liddelow, Marsh, & Stevens, 2020). The second challenge is to analyze a sufficiently large number of subjects to ensure that patient heterogeneity, a critical factor for drug development (Devi & Scheltens, 2018) manifests itself. The studies performed by (B. Zhang et al., 2013) and (Neff, 2021) analyzed 1647 and 364 subjects, respectively, whereas (Mathys et al., 2019) and (Grubman et al., 2019) examined only 48 and 12 subjects, including patients and controls. Only (Neff, 2021) reports molecular heterogeneity among individuals with same clinical diagnosis, but astrocytic data are scarce.

The main goal of this study was to extract consensus astrocytic data from bulk transcriptomic data from three large independent clinical AD databases, encompassing 766 subjects. ‘Consensus data’ was defined as data detected in at least two out of the three databases. Identification of astrocyte-specific genes was optimized by taking a reverse approach as compared to previous studies: instead of determining *a posteriori* which self-organized gene modules might be astrocytic, we localized pre-determined astrocytic gene clusters in whole-brain AD transcriptomes with two-dimensional hierarchical clustering. The workflow is in **Fig. 1**. First, we classified brain genes as cell-specific or non-specific using RNAseq data from astrocytes, neurons, microglia, endothelial cells and oligodendrocytes isolated from aged healthy human brains (Y. Zhang et al., 2016), and a combination of two algorithms: hierarchical clustering, and a recently proposed univariate cell-type enrichment τ scoring (Kryuchkova-Mostacci & Robinson-Rechavi, 2017). Second, astrocytic and, for comparison, neuronal genes were manually annotated according to functions, including cell-type specific functions. Third, the gene clusters were localized in three AD whole-brain transcriptomes generated by the Mount Sinai Hospital (MtSINAI) (B. Zhang et al., 2013) the Mayo Clinic (MAYO) (Allen et al., 2016), and the Religious Order Study and the Rush Memory and Aging project (ROSMAP), which also contains subjects diagnosed with MCI (Bennett et al., 2018). These databases present the advantages of including large cohorts (78-219 subjects), which greatly facilitates the use of multivariate systems approaches, and of having been generated by two different approaches, microarrays and RNAseq, thus diminishing technique-associated bias. Finally, alteration of astrocytic and neuronal functional categories in AD and MCI groups versus controls was statistically established using gene set variation analysis (GSVA), and principal component analysis (PCA). We present the most comprehensive transcriptomic analysis of human AD astrocytes to date using three independent cohorts. As advocated in a recent consensus article about reactive astrocytes (Escartin et al., 2021), the impact of pathway alterations suggested by omics was interpreted in the context of astrocyte biology, instead of resorting to simplistic categorizations of astrocytes as ‘neuroprotective’ or ‘neurotoxic’ from the point of view of neurons. In this vein, our analysis points to dysfunction of the endolysosomal system/mitochondrion axis as a key driver of the phenotypical transformation of reactive astrocytes in AD.

**Figure 1.**
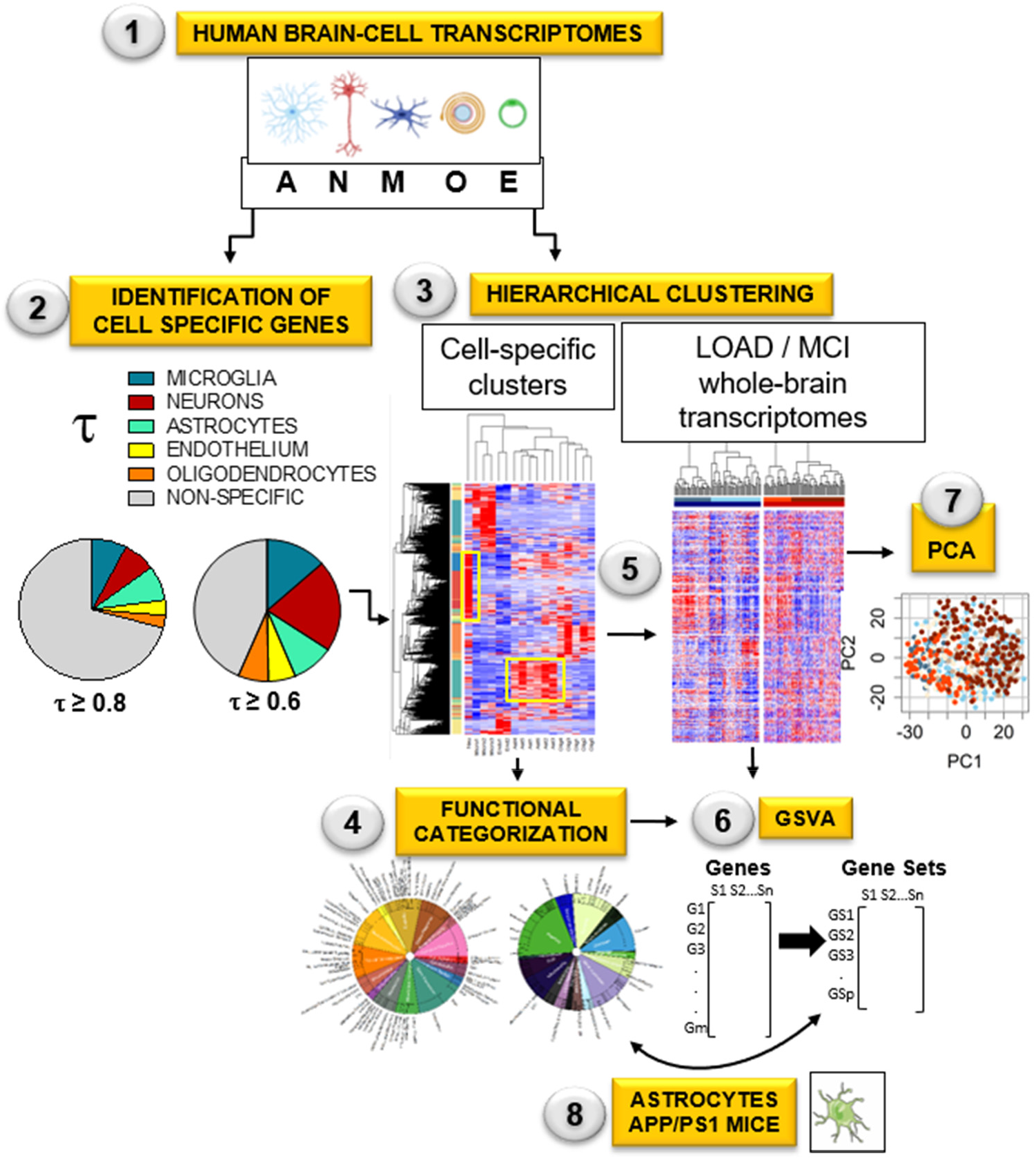
Workflow for the identification of altered astrocytic functions in AD and MCI. (See Methods for details). *Step1*: Genes from human databases were prepared to be classified as cell-specific or enriched using RNAseq data from cells isolated from aged healthy human brains using the univariate cell-type enrichment τ scoring (*Step 2*) and hierarchical clustering (*Step 3*). *Step 4*: The astrocytic and neuronal gene clusters were manually categorized. *Step 5*: The clusters were localized in control, AD and MCI whole-brain transcriptomes from the MtSINAI, MAYO and ROSMAP databases. *Step 6*: Alterations of astrocytic and neuronal functional categories in AD and MCI groups versus controls was statistically established using (*Step 6*) gene set variation analysis (GSVA), and (*Step 7*) principal component analysis (PCA). *Step 8*: To compare patients with mice, the published transcriptome of APP/PS1 astrocytes was re-analyzed using GSVA and our functional categorization and GSVA. The figure in Step 1 was created using cell art adapted from Servier Medical Art (https://smart.servier.com/).

## Material & Methods

### Transcriptome datasets

The cell-type specific RNAseq datasets generated from healthy human brains (Y. Zhang et al., 2016) were downloaded from the National Center for Biotechnology Information (NCBI) Gene Expression Omnibus (GEO) under accession number GSE73721 pre-aligned to gene symbols by the authors using human genome version 19. The database consisted of transcriptomic data from brain cells isolated from temporal cortical lobes of juvenile (8-18 years old) and adult (21-63 years old) non-demented individuals. Transcriptomes were included from 12 astrocytic, one neuronal, five oligodendrocytic, three microglial, and two endothelial samples from individual patients (**Supplementary file 1**, tab 2). Principal component analysis (PCA, computed in R, R Foundation for Statistical Computing, Vienna, Austria) revealed that transcriptomic data of astrocytes isolated from young (<47 years old) and old (>47 years old) were separated along principal component 1 (PC1, **Supplementary file 1**, tab 3). Since the goal of the study was to identify cell-specific signatures in aged subjects, data from young individuals were discarded, retaining a total of six astrocyte transcriptomes from 47–63-year-old subjects, together with all 11 transcriptomes from the remaining cell types. A threshold was set for gene expression such that a given transcript should present a count > 1 in reads per kilo base per million mapped reads (RPKM) in at least one sample. Genes with no counts > 1 were discarded from the analysis. 11077 genes were above this threshold (**Supplementary file 1**, tab 4).

The MtSINAI database containing microarray data of samples from the prefrontal cortex of 101 controls and 129 AD patients obtained from the Harvard Brain Tissue Resource Center was downloaded from GEO under accession number GSE44770 (B. Zhang et al., 2013). The Rosetta gene identifiers were converted to the corresponding gene symbols using the Rosetta/Merck Human 44k 1.1 microarray platform table under accession number GPL4372. When multiple Rosetta identifiers mapped to the same gene, the identifier with the greatest variance across samples was selected. The MAYO database, which contains data from the temporal cortex of 78 control samples and 82 AD samples, was downloaded from Synapse.org under Synapse ID syn3163039 (Allen et al., 2016). The data were downloaded as MayoRNAseq_RNAseq_TCXCounts_normalized.tsv, pre-aligned to Ensemble Gene Identifier. Ensemble Gene IDs were then converted to HGCN gene symbol using `biomaRt` v. 2.32.1 in R (r-project.org). When multiple Ensemble Gene IDs mapped to the same gene symbol, the one with greatest variance was selected. The ROSMAP database (Bennett et al., 2018) contains data from the dorsolateral prefrontal cortex obtained from autopsied non-demented individuals or patients diagnosed with AD. The data were downloaded from Synapse.org under synapse ID syn8456629 as ROSMAP_DLPFC_netResidualExpression.tsv. The dataset contained 200 control cases, 157 MCI cases, and 219 AD cases. In the MCI group, there was no stratification into MCI and MCI-AD. The dataset as posted by the authors had been adjusted via voom (mean-variance modelling at the observational level) normalization to remove bias associated with batch number, RNA integrity, and other data acquisition and processing co-variates. Missing transcript values in the dataset were imputed using a nearest neighbor averaging method using the `impute` package in R.

### Univariate cell-type enrichment score

To determine the cell-specificity of human genes listed in the brain-cell transcriptomics database (Y. Zhang et al., 2016), we used the τ method, reported to be particularly robust compared with other scoring approaches to identify tissue specificity of a gene among different magnitudes of expression and sizes of datasets (Kryuchkova-Mostacci & Robinson-Rechavi, 2017). The τ enrichment score was computed as:

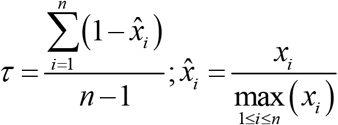

where *x*_*i*_ is the mean expression of the gene within *i*^*th*^ cell type and *n* is the number of cell types. We then define the cell type enriched in the gene as that with the greatest average expression among all cell types. Top τ scores per gene are in **Supplementary file 1**, tab 5. A score of τ ≥ 0.8 indicates that the gene is specific for a given cell type, and a score of 0.8 > τ ≥ 0.6 that the gene is enriched in that cell type. For simplicity, henceforth we refer to genes with τ ≥ 0.6 as ‘enriched’, although note that the ‘enriched’ pool also includes specific genes. Finally, τ < 0.6 indicates that the gene is non-specific, although it is assigned to the cell type with the greatest average expression.

### Hierarchical clustering

The cell type specific data from (Y. Zhang et al., 2016) (**Supplementary file 1**, tab 4) were z-scored for each gene across samples. Gene and sample hierarchical clustering were conducted using a Euclidian distance measure, and the average agglomeration method in R. Gene clusters associated with neurons and astrocytes were determined by cutting the dendrogram at a height of 3.76 to achieve that at least 70% of the genes in the neuron and astrocyte clusters have the highest τ for that cell type (results from hierarchical clustering are in **Supplementary file 1**, tabs 6-8). Corresponding cell-specific gene clusters in whole tissue data sets (MtSINAI, MAYO, and ROSMAP) were identified by sorting the genes of whole-tissue data according to cell type specific datasets. Whole-tissue subject samples (columns) were then clustered within each Control, MCI, or AD cohort (Euclidian distance and Ward.D2 agglomeration) to identify molecularly-defined sub-groups of samples.

### Data normalization within each cell type

A recognized caveat of bulk transcriptomic analyses is that DEG may reflect changes in cell-type composition rather than real changes in cell-specific transcriptomes. This is relevant to AD because there is neuronal death (Gomez-Isla et al., 1997), and microglia proliferation (Marlatt et al., 2014). Since neurons are the most predominant cell type in the brain (40% in the human neocortex (Pelvig, Pakkenberg, Regeur, Oster, & Pakkenberg, 2003)), a decrease of the neuronal pool, with no change in the astrocyte pool (Marlatt et al., 2014), arguably results in the artificial up-regulation of astrocyte genes. Several approaches have been employed to correct for cellular composition, such as use of several of the so-called ‘high fidelity’ genes primarily expressed by one cell type (Kelley, Nakao-Inoue, Molofsky, & Oldham, 2018). Here, we reasoned that reliance on a subset of highly cell-specific genes could lead to artifacts should their expression change in the course of the disease, because there is no assurance that they are stably expressed as house-keeping genes. Therefore, we elected to remove gross variance by normalizing gene expression within the neuronal and astrocyte clusters according to the shifts in distribution of differential expressions of all genes. This was accomplished by variance stabilizing normalization using `limma` in R, which simultaneously correct each dataset for dependence of variance on the mean. Removal of gross variance is widely-used in normalizations of transcriptomic data, based on the premise that, since only a negligible proportion of genes have their expression changed following perturbations (Lin, Du, Huber, & Kibbe, 2008), a bulk shift in the gene-expression distribution in a given cell population reflects changes in the proportion of that cell type among brain cell populations. Thus, correcting gene expression according to the shift would unmask the real changes in gene expression. We applied this correction separately to the neuronal and astrocytic gene clusters.

### Cell function annotation and gene set variation analysis

Neuronal and astrocytic gene clusters generated by combining τ (Kryuchkova-Mostacci & Robinson-Rechavi, 2017) and hierarchical clustering of transcriptomes from cells from healthy brains (Y. Zhang et al., 2016), were organized in functional categories and subcategories (**Supplementary file 2**). Categories were manually curated by one expert and cross-validated by another expert using information from GeneCards, perusal of Medline and PubMed, and open-source platforms such as GO, KEGG, and Reactome. Manual curation was indispensable for two reasons. First, over 60% of the genes in the astrocytic cluster were not annotated in open-source platforms. This means a substantial loss of usable gene data if we only relied on genes annotated in open-source platforms. Second, annotation in such platforms represents ‘canonical’ pathways with little consideration for the fact that different CNS cell types are molecularly and functionally distinct (Y. Zhang et al., 2016). This limitation is critical for astrocytes. For example, unlike neuronal compartments such as ‘synapses’, ‘dendrites’ and ‘spines’, a highly specialized astrocytic compartment termed ‘perisynaptic astrocyte processes’ (PAP), concentrating RNA transcripts related to glutamate and GABA metabolism, energy metabolism, as well as the ribosomal machinery to perform local RNA translation (Sakers et al., 2017), is not a category in current KEGG and GO databases. The genes were organized in general categories and subcategories using a mixed set of criteria including general pathways (e.g. carbohydrate metabolism or amino acid metabolism) and subcellular compartments with a clear functional specialization (e.g. mitochondria, peroxisome or lysosome). Such flexibility allowed us to optimize gene inclusion, while generating gene sets with sufficient numbers of genes for statistical purposes. Although most genes were assigned to only one category, around 100 genes were assigned to two categories (in blue, **Supplementary file 2**, tab ‘astrocyte cluster 4’). The majority of those genes (96) were PAP genes, because we reasoned that their presence in PAP does not exclude relevance in other subcellular compartments. For example, *ALDOC* and *ACO2* are, plausibly, ubiquitous enzymes relevant for general glycolysis and mitochondrial Tricarboxylic Acid (TCA) cycle throughout the cell. Likewise, genes related to ‘TNFalpha signaling’ are in ‘stress responses’/’cytokines’ as well as in ‘gliotransmission’, for TNFalpha has been described to modulate glutamate exocytosis (Santello, Bezzi, & Volterra, 2011). Overlap between groups was less than 5% with the exception of ‘PAP and gliotransmission’: 17.8% genes in ‘gliotransmission’ were in ‘PAP’, and 10.3% of PAP genes were in ‘gliotransmission’. For neurons, each gene was assigned to only one category.

Using our novel annotation database, the `GSVA` package in R was used to identify the enrichment of each gene set across all samples. GSVA is a generalized gene set enrichment method that detects variations of pathway activity over a sample population in an unsupervised manner (Hanzelmann, Castelo, & Guinney, 2013). Statistical differences in enrichment scores for each gene set between subject groups were computed by comparing the true differences in means against a null distribution obtained by permuting the gene labels and re-computing the GSVA 1000 times. False discovery rate adjust p-values (qFDR) were computed using the method of Benjamini & Hochberg (Hanzelmann et al., 2013). Gene sets with FDR adjusted p value qFDR < 0.05 were considered significant.

### Relationships between clinical stages and GSVA scores

The ROSMAP dataset includes Braak and CERAD (Consortium to Establish a Registry for Alzheimer’s disease) pathological scores, APOE genotype, and sex for each sample. To determine the relationship between these phenotypes and the GSVA enrichment scores for each sample, we used the `stats` R package to fit linear models between enrichment scores and either Braak or CERAD for each gene set. The MAYO and MtSINAI databases do not include Supplementary information per subject other than the clinical classification.

## Results

### Hierarchical clustering identifies groups of cell-type specific genes

To identify astrocyte-specific genes, we calculated the metric τ (Kryuchkova-Mostacci & Robinson-Rechavi, 2017) for every gene in the transcriptomes from isolated brain cell types generated by Zhang et al. 2016 (**Supplementary file 1**, tab 5 ‘τ scores’). Cell type-specific (τ ≥ 0.8) and cell type-enriched (τ ≥ 0.6) genes represented 30% and 60% of the 11,077 genes, respectively. Astrocytes (8%), microglia (7.6%) and neurons (7.2%) present the largest percentage of cell-specific genes, whereas neurons (21%) contained the largest pool of cell-enriched genes, followed by microglia (13.4%) and astrocytes (9.4%). In contrast, endothelial cells and oligodendrocytes presented the lowest percentages of cell-enriched (6%) and specific (3%) genes.

In parallel, because cell-specific functions are arguably performed by co-expressed genes (Weirauch, 2011), we reasoned that cell-type specific genes would be identified by clustering analysis. Thus, we applied hierarchical clustering (**Methods**) to the cell-type specific dataset, generating 194 clusters that contained between 1-1651 genes (**Fig. 2A**; **Supplementary file 1**, tab 6 ‘Hierarchical clustering’ and tab 7 ‘Cluster description’). We found that genes from the same cell type segregated together, defining clusters of highly co-expressed genes, plausibly representing cell-specific functions. The dendrogram height of hierarchical clustering was set to identify gene clusters that were highly enriched for individual brain cell types. The criteria for ‘enrichment’ was that the cluster contained more than 100 genes, of which over 70% corresponded to a unique cell type with τ ≥ 0.6 (**Supplementary file 1**, tab 8 ‘cluster selection’). Five clusters fulfilled these criteria, corresponding to astrocytes (cluster # 4), endothelial cells (cluster # 5), neurons (cluster # 13), microglia (cluster # 17) and oligodendrocytes (cluster # 164). There were other clusters containing over 77% of total of neuronal (e.g., clusters # 10, 116) or microglial (e.g., clusters # 30, 15) genes (**Supplementary file 1**, tab 8 ‘cluster selection’), but they were excluded from the analysis because less than 54% of their genes was enriched (τ ≥ 0.6) for that cell type, suggesting lower cell type specificity.

**Figure 2.**
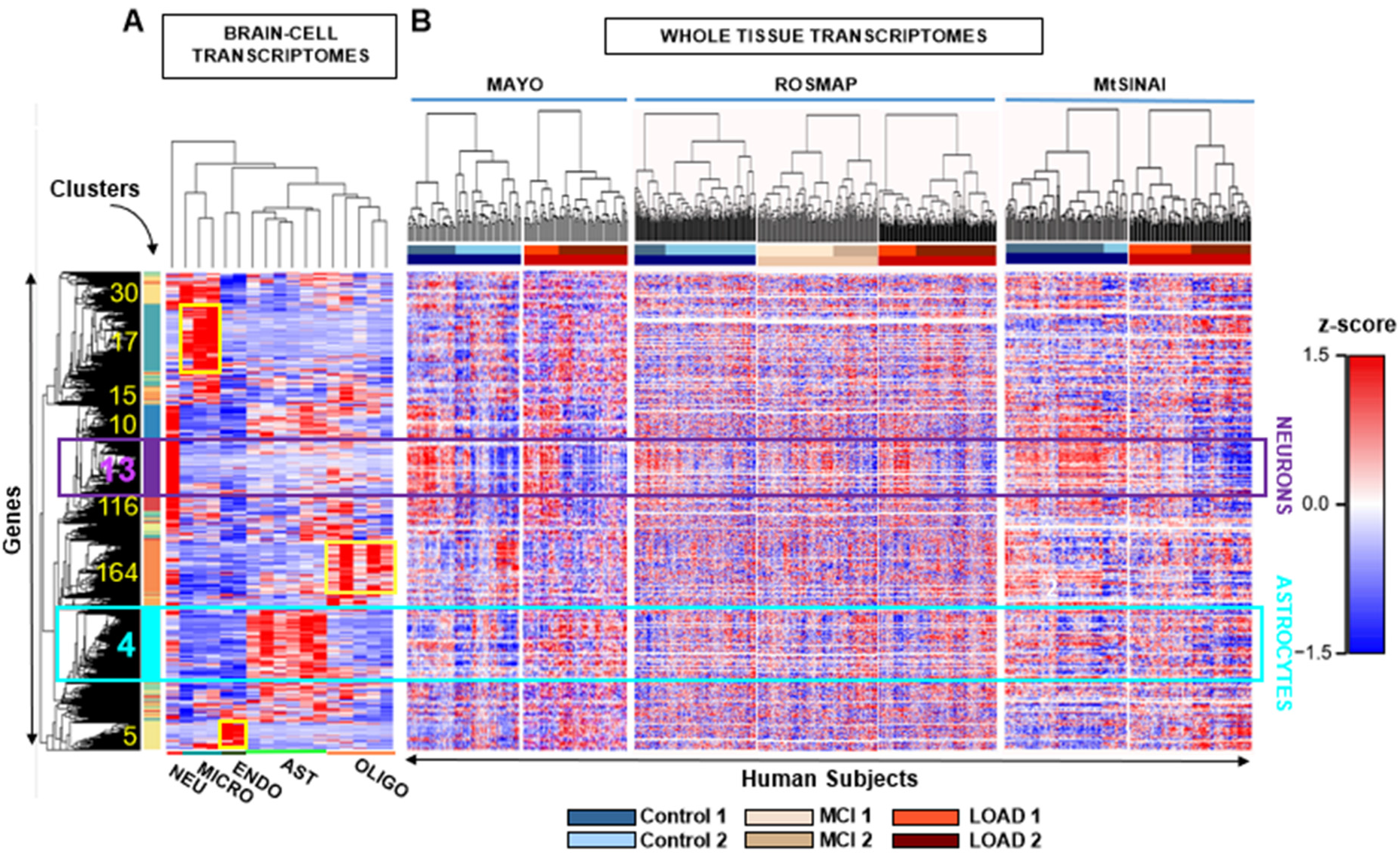
Identification of cell-specific gene clusters in whole-brain transcriptomes by two-step hierarchical clustering. **A**. Hierarchical clustering was performed on transcriptomes of brain cell populations isolated from individual healthy subjects. Each column is a sample from a single subject/patient and each row is a z-scored gene. In yellow, the number and location of clusters with highest z scores, coinciding with highest τ scores for a given cell type (cluster information in **Supplementary file 1**, tabs 6-8). The turquoise and purple squares are the main astrocytic and neuronal clusters (cluster # 4 and cluster # 13), respectively. Microglia, oligodendrocyte and endothelial-cell clusters are 17, 164 and 5, respectively (shown in yellow). Clusters 30, 15, 10 and 116 are highly enriched in microglial or neuronal genes, but fall below the criteria established for cell-specificity (**Methods**). **B**. Whole-brain transcriptomes from individual subjects of control (blue), MCI (beige) and AD (red) groups from MAYO, ROSMAP, MtSINAI were hierarchically clustered. The rows from the cell-type clustering in **A**. were kept fixed to identify cell-specific clusters. The figure was generated with raw data before correction for cell composition. Clustering of subjects segregated all cohorts into at least 2 subgroups (defined as 1 and 2) showing opposite changes in gene-expression.

We continued with astrocyte cluster 4 (1651 genes) and neuronal cluster 13 (1258 genes) for two reasons: (i) a cluster size of over 1000 genes is sufficient for functional categorization and gene set analysis and (ii) since molecular changes in neurons are better characterized in AD than changes in astrocytes, neurons served as a positive control to assure correct data mining.

In the neuronal cluster # 13, 99.9 % of the genes were neuronal according to τ score, of which 54.1% of genes were neuron-specific (τ ≥0.8), meaning that they encode for proteins that perform functions highly specific to neurons. Indeed, the top 10 ranked genes based on τ (τ > 0.98) were related to neurotransmission (*SYNPR, GABRA1, CNR1, SYT1, GABRG2* and *GAD1)*, regulation of membrane potential (*KCN2*), microtubules (*INA)*, and cell adhesion *(RELN*). The remaining 45.9% (τ < 0.8) were neuron-enriched or bulk genes plausibly supporting neuron-specific functions, since they were co-expressed with neuron specific genes (**Supplementary file 1**, tab 8 ‘cluster selection’; **Supplementary file 2**, tab 2 ‘Neurons cluster 13’).

In astrocyte cluster # 4, 80% of the genes were astrocytic according to τ (**Supplementary file 1**, tabs 7 and 8 ‘Cluster description’ and ‘Cluster selection’; **Supplementary file 2**, tab ‘Astrocytes cluster 4’). Further, 30% had τ ≥ 0.8, including genes that encode typical astrocyte proteins such as *GFAP* (τ = 0.95), glutamine synthase (*GLUL*, τ = 0.95), glutamate decarboxylase (*GLUD2*, τ = 0.91), glutamate transporters (*EAAT1/SLC1A3*, τ = 0.83), lactate dehydrogenase (*LDHB*, τ = 0.91), and enzymes involved in mitochondrial fatty-acid oxidation (ACADVL, τ = 0.92). Enriched genes with τ between 0.6-0.8, or bulk genes below 0.6, arguably necessary for astrocyte functions, are the aquaporins A*QP1* (τ = 0.739) and *AQP4* (τ = 0.744), and the chaperone clusterin *CLU* (τ =0.373), which was identified by GWAS as a risk factor in AD (Han, Qu, Zhao, & Zou, 2018). These findings indicated that the group of genes gathered in cluster #4 is relevant to astrocyte biology.

### Identification of cell-type specific gene clusters in AD databases

We next searched for the astrocytic and neuronal gene clusters in gene expression data from three AD whole-tissue transcriptomic databases (heatmaps of z-scores in **Fig. 2B)**. Unexpectedly, hierarchical clustering of subjects (columns) of each cohort revealed heterogeneity in groups with the same clinical diagnosis. Subjects were divided in at least two molecular groups within the Control, MCI, and AD cohorts across the three databases, as shown by the division of the dendrograms into two large groups in each cohort. We refer to *type 2* subjects as those showing blue z-scores within the neuron gene cluster, suggesting massive down-regulation of genes, as compared to *type 1* subjects, in which genes with red z-scores predominated. Type 1 sub-cohorts included Control1, MCI1 and AD1, while *type 2* included Control2, MCI2 and AD2. Numbers of Control1/Control2 patients were 33/45 in Mayo, 51/149 in ROSMAP, and 81/20 in MtSINAI. Numbers of AD1/AD2 patients were 28/54 in Mayo, 70/149 in ROSMAP, and 66/63 in MtSINAI. MCI1/MC2 numbers were 99/58 in ROSMAP. We note that heterogeneity in MtSINAI data has been recently reported and replicated in ROSMAP (Neff, 2021).

The most profound down-regulation of genes within the neuronal gene cluster was observed in AD2. Because down-regulation of neuronal genes due to loss of synapses and neuronal demise is a hallmark of AD (Gomez-Isla et al., 1997), we reasoned that AD2 was advanced AD, and Control1 a *bona fide* control composed of subjects that were neither demented, nor preclinical or pre-symptomatic at the time of death. Control1 and AD2 thus appeared to be the extreme groups across the continuum of AD phenotypes, and were thus selected for further analysis.

Since our goal was to identify changes in astrocytic functions, it was critical to eliminate gross variability among samples within the astrocytic and neuronal gene clusters due to global changes in cell composition. To do so, we applied variance stabilizing normalization (**Methods**), which shifted the peaks of the distributions of differences in gene expression between AD2 and Control1 to zero (**Fig. 3A**). Prior to normalization, the distributions of gene-expression differences shifted to the left in the neuronal cluster, suggesting depletion of neurons in AD2, while the distribution of astrocytic genes shifted to the right, suggesting a relatively higher content of astrocytes in AD2. After normalization, the global downregulation of all neuronal genes in cluster # 13 in *type 2* groups was attenuated, but intra-cohort heterogeneity nevertheless persisted such that type 2 clusters presented prominent down-regulation of neuronal genes as compared to type 1 clusters (**Fig. 3B** and **Supplementary file 3**). In addition, new gene clusters that were previously masked were unraveled by hierarchical clustering. Thus, there were three neuronal (**Fig. 3B**, N-a, b, c), and four astrocytic gene subgroups (**Fig. 3B**, A-a, b, c, d) with opposite expressions patterns in *type 1* versus *type 2* subjects. For example, N-b genes were globally downregulated in AD2 and Control 2, as compared to Control 1 and AD1. Importantly, N-b contained genes related to synaptic function, including glutamatergic and GABAergic neurotransmission (e.g., *GABRB3; GABRB2; GAD1; PDYN; SYN2; GABRG2; SYN1 NAPB; GRIA2; SLC17A7; GRIK2*), supporting that this cohort is a *bona fide* AD. The normalized expression of cluster # 4 genes in the three AD databases is in **Supplementary file 4**.

**Figure 3.**
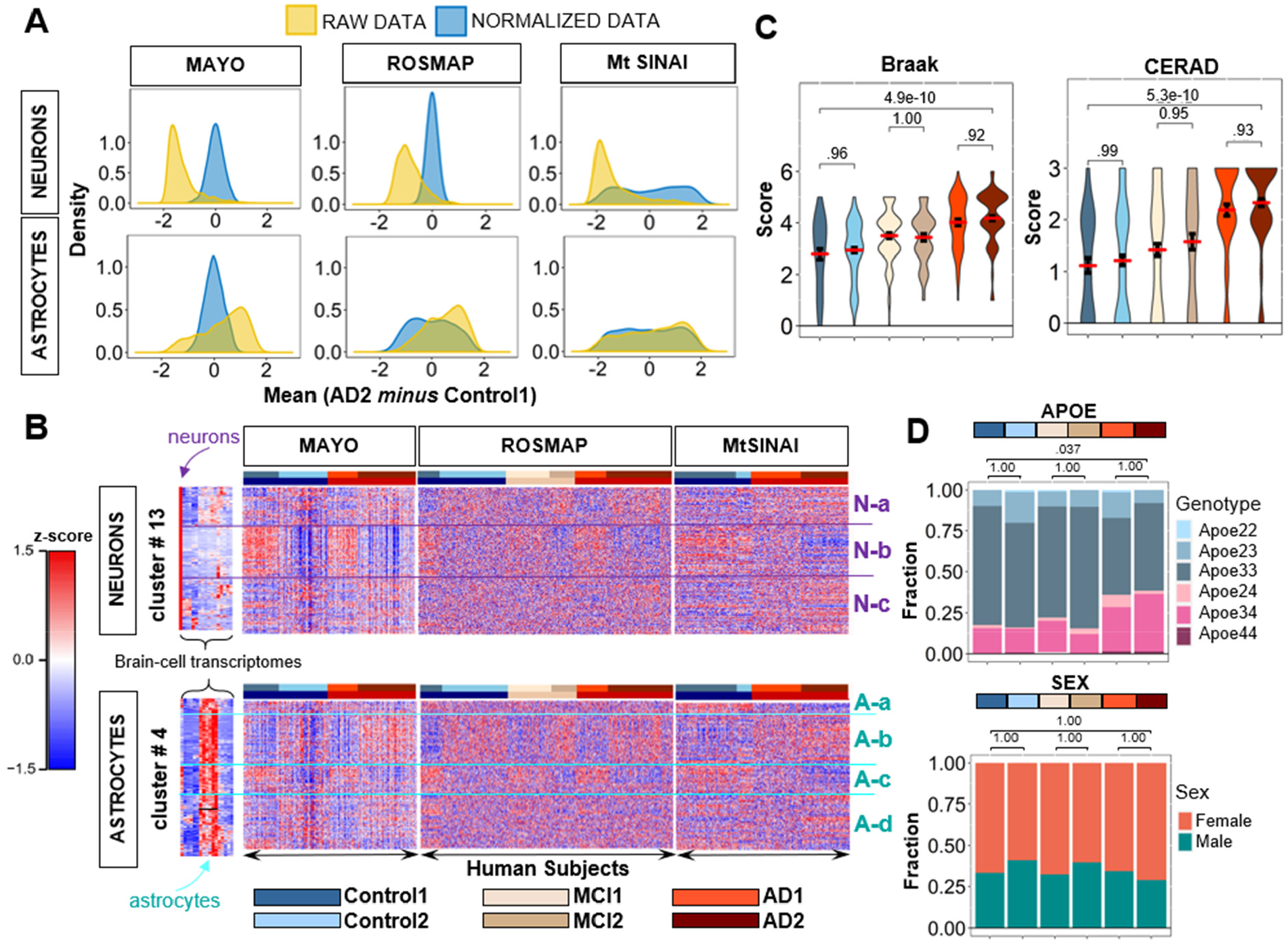
Neuronal and astrocytic gene sub-clusters in the three clinical databases. **A**. Distributions of gene expression differences between AD2 and Control1 cohorts before (yellow) and after (blue) variance stabilizing normalization, yielding zero-centered distributions (**Methods**). **B**. Hierarchical clustering of Control (blue), MCI (beige), and AD (red) subjects from Fig. 2B using normalized data from Fig. 3A in rows. For both astrocytic and neuronal genes, subjects self-organized into at least two distinct subgroups (defined as 1 and 2) within each clinical group, while genes self-organized into up to four groups (Na-c and Aa-d). In MAYO and ROSMAP, clusters showed opposite directions of change in *type 1* and *2* subjects. In MtSINAI, Control2 was like Control1, and AD1 like AD2, although with less pronounced changes. **C**. Violin plots of pathological stages according to CERAD and Braak scoring shows no significant difference in pathological stages between sub-groups within each cohort in ROSMAP (means ± SEM, one-way ANOVA with Tukey post-hoc). **D**. Distribution of *APOE* genotypes and sex in the six groups. While 3/3 was the predominant genotype across all groups, and 3/4 was more abundant in AD, there were no differences in frequency of *APOE4* alleles between *type 1* and *2* subgroups within each clinical group. Likewise, females were more abundant than males in all groups, but not in *type 2* subjects as compared to *type 1* (p values determined by pairwise Fisher’s Exact test with Bonferroni adjustment).

Because the ROSMAP database included clinical covariate data, such as CERAD and Braak scores, APOE genotype and sex, we examined whether *type 1* or *type 2* sub-cohorts were associated with specific covariates to gain insight into why they were molecularly heterogeneous. We found that Braak and CERAD scores were significantly higher in AD groups than in Controls (**Fig. 3C**), confirming the clinical diagnosis. However, *type 1* and *type 2* subjects with the same clinical diagnoses did not significantly differ in their pathological stages according to CERAD and Braak scoring; nor were type 2 groups more enriched in *APOE4* subjects or females (**Fig. 3D**).

#### GSVA reveals altered functions in AD and MCI versus Control1

We next identified changes in astrocytic and neuronal functions using the normalized expression data. Because the majority of the astrocytic genes were not annotated in existing pathways in open-source databases, we manually curated the astrocytic cluster in 17 functional categories (gene sets) and 135 subcategories, and the neuronal cluster in 15 functional categories and 79 subcategories (**Methods, Supplementary file 2**).

GSVA was then used to compare the gene sets corresponding to different functional categories among cohorts, yielding an enrichment score for each subject and gene set. We started by comparing the extreme cases, AD2 and Control1, using two criteria. First, probability values were computed using a permutation analysis across genes (cut-off for statistical significance was set at FDR-adjusted q < 0.05, **Methods**). Second, we considered that a function was altered in AD2 as compared to Control1 if such alteration was detected in at least two out of the three databases (consensus criterion).

For neurons, seven functions were altered in AD2 versus Control1 **(Fig. 4A**, statistics in **Supplementary file 5**, extended graphs in **Supplementary file 6**): ‘neurotransmission’ and ‘gene expression’ were altered in the three databases, and ‘synaptic plasticity’, ‘regulation of membrane potential’, ‘neural development’, ‘mitochondria’ and ‘stress response’ in MAYO and ROSMAP, but not MtSINAI. ‘Synaptic plasticity’, ‘neurotransmission’, ‘neural development’ and ‘membrane potential’ were down-regulated, and ‘gene expression’, ‘mitochondria’ and ‘stress response’ up-regulated. Among significantly different gene sets in MAYO and ROSMAP (**Fig. 4A**), Control 2 and MCI2 resembled AD2, and AD1 and MCI1 resembled Control1 (**Fig. 4B**). In MtSINAI, we found no statistically significant difference between Control 1 and Control 2, and AD groups were similar, although changes in AD2 were more dramatic, as compared to Control1 (**Fig. 4B**).

**Figure 4.**
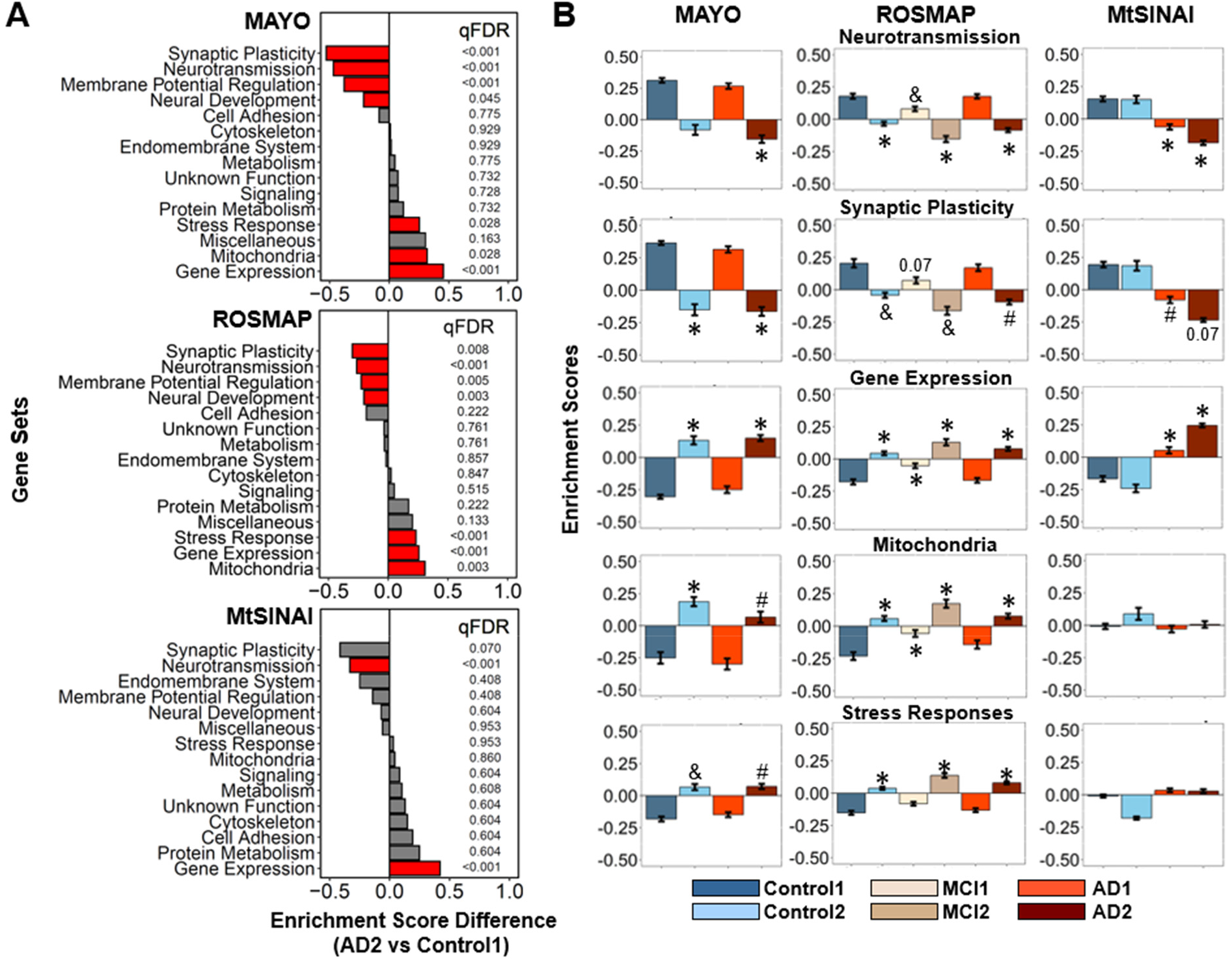
GSVA unravels neuron-enriched functions altered in AD. Differences in the general functional categories among cohorts in MAYO, ROSMAP and MtSINAI databases were examined by GSVA. **A**. Comparison between AD2 and Control1. Functions are ranked according to the difference in enrichment scores (ES). Significantly changed functions (qFDR < 0.05) are labeled in red. **B**. Enrichment scores of functional categories significantly changed in at least 2 out of 3 databases. (#) qFDR< 0.05 (&) qFDR <0.01, and (*) qFDR < 0.001, versus Control1.

For astrocytes, five functions were altered in AD2 versus Control 1 (**Fig. 5A**, statistics in **Supplementary File 5**): ‘PAP’, ‘plasticity’ ‘stress response’ were up-regulated in three/three databases, ‘mitochondria’ was down-regulated in three/three databases, while ‘endomembrane system’ was down-regulated in two/three databases. Because ‘PAP’ are specialized astrocyte-neuron contacts and, hence, this category includes many genes related to gliotransmission, and since ‘gliotransmission’ was highly significantly changed in ROSMAP (qFDR<0.001), and trending in MAYO (qFDR=0.07), henceforth ‘PAP’ and ‘gliotransmission’ were considered together. As with neurons, Control2 and MCI2 showed the same direction of change as AD2 compared to Control1 (**Fig. 5B**). Likewise, enrichment of these gene sets in AD1 resembled Control1, particularly in MAYO and ROSMAP, while AD1 appeared to be a milder version of AD2 in MtSINAI.

**Figure 5.**
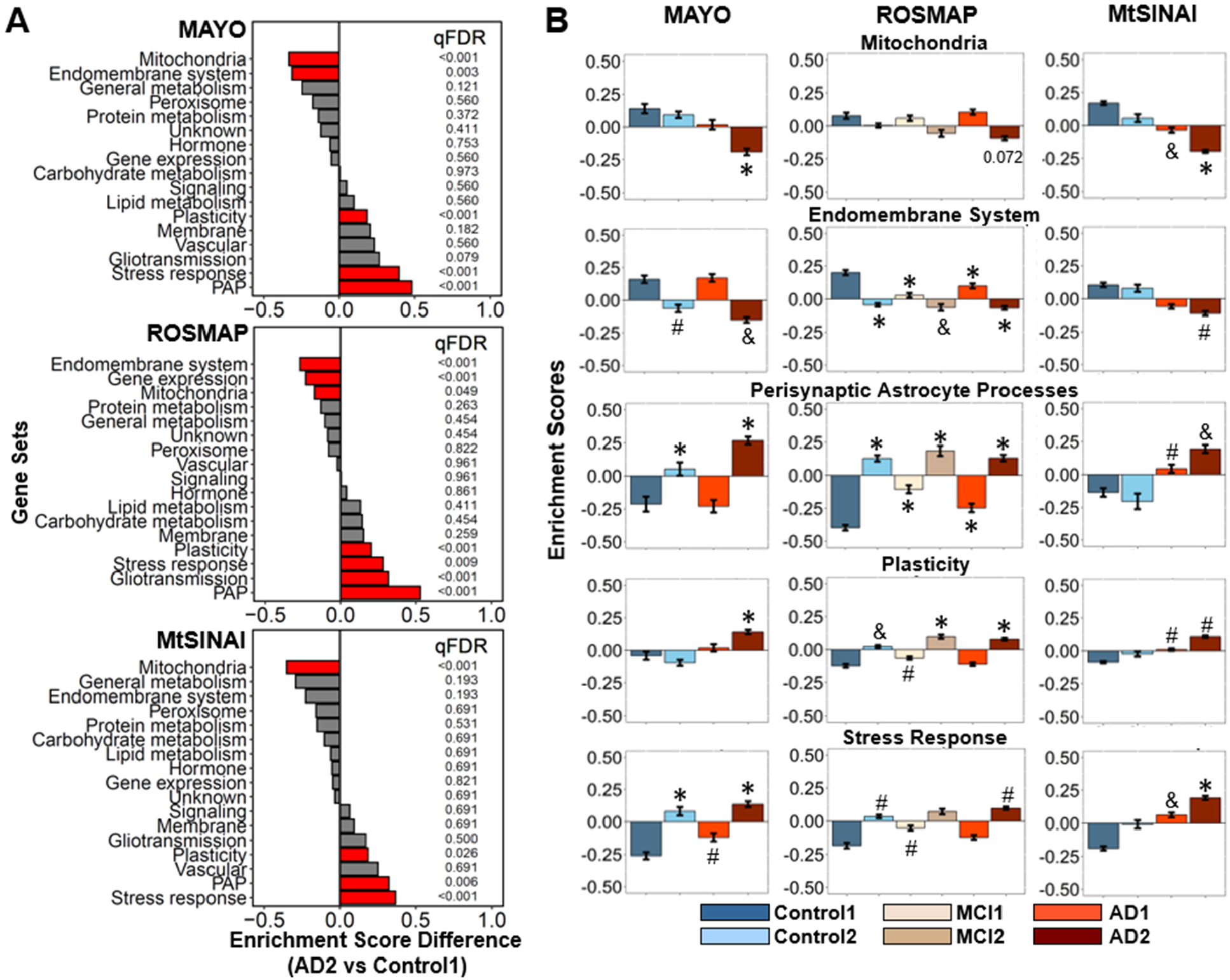
GSVA unravels astrocyte-enriched functions altered in AD. Differences in the functional categories among cohorts in MAYO, ROSMAP and MtSINAI databases were examined by GSVA. **A**. Comparison between AD2 and Control1. Functions are ranked according to the difference in enrichment scores (ES). Significantly changed functions (qFDR < 0.05) are labeled in red. **B**. Enrichment scores of functional categories significantly changed in at least 2 out of 3 databases. (#) qFDR < 0.05, (&) qFDR <0.01, and (*) qFDR < 0.001, versus Control1.

### Co-variance of gene sets with one another and with pathological covariates

#### Correlations between altered functions and pathological stages

Since CERAD and Braak scores were available for the ROSMAP data, we examined relationships between the enrichment of each gene set and pathological stages by regressing enrichment scores for each gene set and sample against the corresponding CERAD and Braak scores, regardless of clinical diagnoses (**Methods**). Data from all the individuals in the three groups (control, MCI and AD) were used in the analysis. For neurons, we found no correlation between Braak and CERAD stages for any of the seven top dysregulated categories described in **Fig. 4A**. The p values for regressions with Braak/CERAD scores were: ‘neurotransmission’ (0.536/0.919), ‘synaptic plasticity’ (0.342/0.703), ‘neural development’ (0.85/0.552), ‘membrane potential’ (0.794/0.479), ‘gene expression’ (0.55/0.563), ‘mitochondria’ (0.357/0.254) and ‘stress response’ (0.548/0.755) **(Supplementary file 7)**. The lack of correlation can be interpreted to indicate similar gene dysregulation in all the stages, suggesting that neuronal dysfunction is an early event in AD pathology such that it appears in subjects with no (i.e., Control2) or incipient (i.e., MCI2) cognitive deficits, and low CERAD and Braak scores.

For astrocytes, of the five functions that were significantly altered in AD2 vs Control1 (**Fig. 5A**), the following ones significantly correlated with Break/CERAD according to p-values: ‘mitochondria’ (0.0000203/0.000742), ‘stress response’ and ‘plasticity’ (0.0768/0.00967) (**Supplementary file 8**). In contrast, ‘PAP’ (0.563/0.838), ‘gliotransmission’ (0.811/0.805), and ‘endomembrane system’ (0.709/0.942) did not correlate with pathological stages, suggesting that, as we reasoned with neurons, they represent early manifestations of alterations in astrocytic functions. For example, there is a progressive decline in the expression of mitochondrial genes from CERAD 1 to 3 and from Braak 1 to 6, while the expression of genes involved in ‘endomembrane system’ is altered even before the detection of cognitive deficits in patients.

#### Correlations among altered functions

We examined correlations among all neuronal and astrocytic gene sets by regressing enrichment scores for each gene set against each other gene set. Hierarchical clustering of correlation coefficients and adjusted p-values are in **Supplementary file 9** for ROSMAP, MAYO, and MtSINAI. In neurons, the strongest direct correlations were among highly related functions such as ‘membrane potential’, ‘neurotransmission’ and ‘synaptic plasticity’ in neurons. Likewise, in astrocytes, the strongest correlations were between ‘PAP’ and ‘gliotransmission’ (orange squares). The strongest anti-correlations point to intertwined changes between: (i) morphological remodeling of PAP and loss of synaptic functions, and (ii) survival pathways included in ‘plasticity’ in astrocytes (see below) and organelle dysfunction.

### Alteration of functional subcategories in astrocytes

We next aimed to gain further insight into which factors drive the functional alterations in AD astrocytes by searching for functional *subcategories* with statistically significant differences in enrichment scores between AD2 and Control1 (qFDR< 0.05) in at least two out of three of the databases (statistics in **Supplementary file 10**).

The category ‘PAP’ includes different categories related to morphology, metabolism and gene expression. The consensus subcategory significantly upregulated was ‘PAP morphology’ (qFDR <0.001, AD2 vs Control1, in MAYO and ROSMAP). Because PAP are specialized in the functional refinement of adjacent synapses at a micro scale (Sakers et al., 2017), increased expression of PAP-related genes in AD astrocytes may be a compensatory reaction to locally optimize synaptic functions.

Subcategories in ‘Plasticity’ include annotations associated with morphological remodeling (e.g., ‘adherens junction’, ‘ECM’, ‘cytoskeleton’, ‘ciliogenesis’, ‘TGFbeta signaling’, ‘metalloproteases’), and pathways involved in brain development whose role in adult astrocytes is not well understood (e.g., ‘axon outgrowth’, ‘synaptogenesis, ‘WNT signaling’, ‘hippo signaling’, ‘smoothened signaling’, ‘hedgehog signaling’, ‘homeobox signaling’, ‘notch signaling’, ‘FGF signaling’, ‘pluripotency’, and ‘glial cell fate commitment’). Significantly dysregulated pathways were ‘ECM’, upregulated in the three databases, and ‘hippo signaling’ and ‘ciliogenesis’ (i.e., the process of generation of a microtubule-based and centriole-derived cilium whose function in astrocytes is unknown), respectively upregulated and down-regulated in two databases—note that the fact that the general trend of a given general category is towards up-regulation does not preclude that some of its subcategories are down-regulated.

‘Stress responses’ includes ‘AMPK signaling’, ‘antioxidant response’, ‘cell death’, ‘chaperone’, ‘DNA repair’, ‘hypoxia’, ‘complement system’ and ‘cytokines’. No subcategory was dysregulated according to qFDR in at least two databases. However, it is worth noting that ‘TNFalpha’, which is a subcategory of ‘cytokines’ as well as of ‘gliotransmission’, was significantly dysregulated in MAYO and ROSMAP, and trending in MtSINAI (qFDR=0.093).

In ‘endomembrane system’, encompassing genes related to ‘endoplasmic reticulum’, ‘Golgi’, ‘lysosome’ and ‘vesicles’, no single subcategories were significantly dysregulated, while in ‘mitochondria’, ‘ETC’ and ‘TCA cycle’ were significantly downregulated in AD2 versus Control1 in three and two databases, respectively. For ‘ETC’, qFDR=<0.001, <0.001, 0.012, <0.001 in MAYO, ROSMAP and MtSINAI. For ‘TCA cycle’, qFDR=<0.001, 0.39, 0.01 in MAYO, ROSMAP and MtSINAI. All in all, the dysregulated subcategories in astrocytes point to alterations in mitochondrial respiration concomitant to morphological remodeling.

### PCA confirms main functional changes in AD astrocytes

GSVA was dependent on gene sets organized into pre-determined functions. Thus, we also used PCA as an alternative, unsupervised approach to gain independent insight into the predominant functional changes of AD astrocytes. First, we examined whether expression of genes contained in the astrocytic cluster # 4 segregated cohorts in ROSMAP. PCA segregated the Control1, MC1 and AD1 groups to the left, and the Control2, MCI2 and AD2 groups to the right, according to the principal component 1 (PC1, horizontal axis, **Fig. 6A**). Violin plots of PC1 scores per group confirmed this tendency.

**Figure 6.**
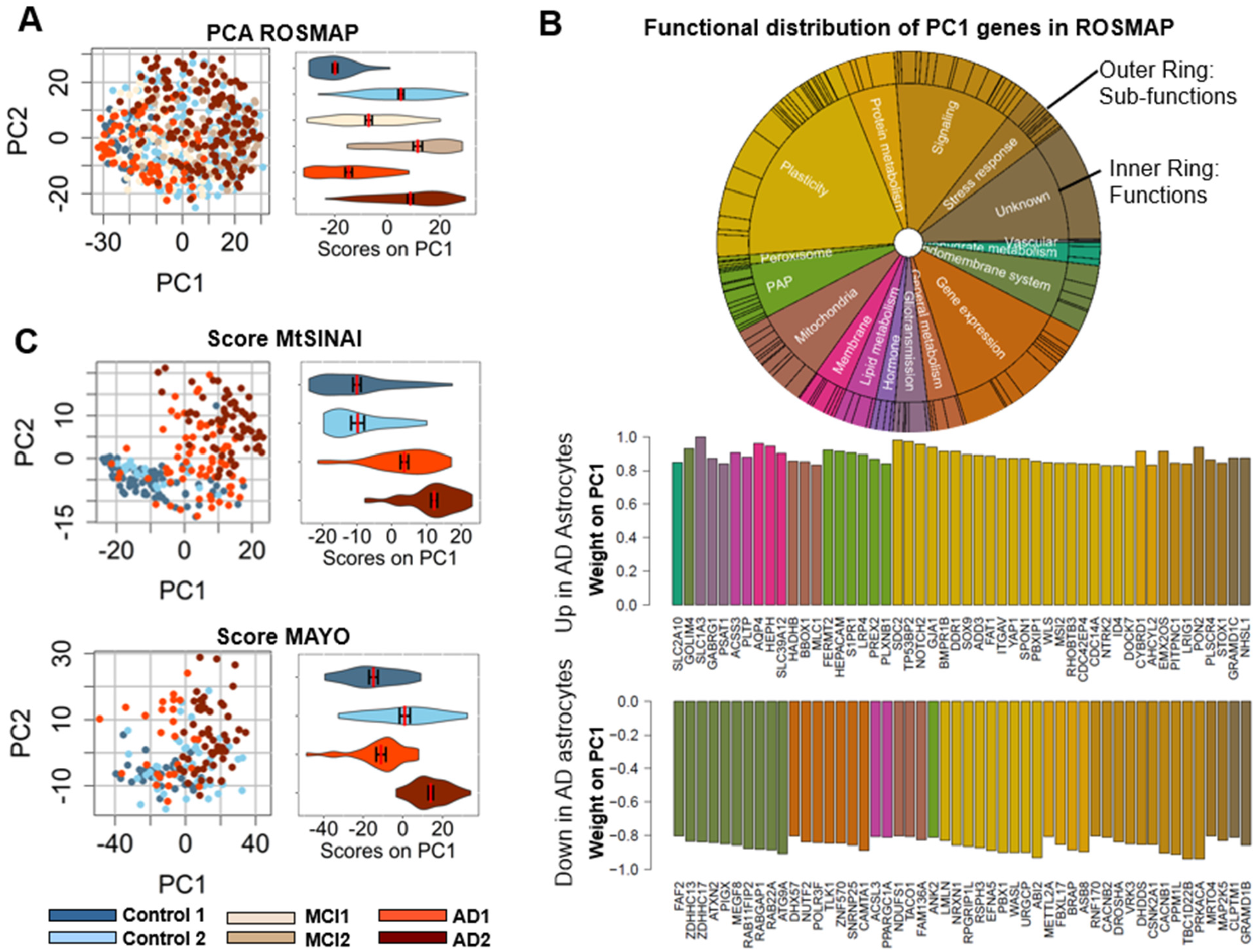
PCA identifies top altered functions in AD astrocytes. **A**. PCA analysis of astrocyte-enriched genes in ROSMAP. Dot plots and violin plots along PC1 (mean ± SEM) show differential expression of astrocyte genes in type 2 versus *type 1* subjects. **B**. Top varying genes in PC1 (up-regulated and down-regulated), color-coded for function according to the circos plot, inform about functions altered in AD astrocytes. The vertical axis represents the relative contribution (importance/weight) of each gene for the scoring of individual samples in panel **A** along PC1. **C**. Scoring of MtSINAI and MAYO databases using genes overlapping with ROSMAP PCA in **A**. discriminates Control and AD cohorts along PC1, as shown by the dot and violin plots (means ± SEM).

Second, we examined the functions performed by the proteins encoded by the top 50 dysregulated genes in PC1, representing the maximally co-varying astrocytic genes (**Supplementary file 11**). Mirroring our results with GSVA, genes related to ‘plasticity’ and ‘PAP/gliotransmission’ were over-represented in the up-regulated portion, whereas genes related to ‘endomembrane system’, ‘gene expression’, and ‘signaling’ were predominant in the down-regulated portion (**Fig. 6B**). Genes related to ‘mitochondria’ and ‘stress responses’ appeared in both sections, indicating mixed direction of dysregulation in these functions. Unlike GSVA, PCA also detected changes in ‘signaling’, as suggested by the presence of genes related to this function in the up and down segments of the top PC1 genes.

Third, we asked if the changes detected in AD astrocytes in ROSMAP with PCA were reproducible. Thus, we determined the capacity of the PC1 genes to segregate cohorts in MAYO and MtSINAI. As in ROSMAP, Control1 subjects were mostly negative, and AD2 cases were mostly positive in MAYO and MtSINAI, thus validating the discriminating capacity of the maximally co-varying genes in PC1 (**Fig. 6C**). AD1 segregated with Control 1 in MAYO, but not in MtSINAI, where AD1 aligned with AD2, confirming less intra-cohort heterogeneity in MtSINAI than in the other two databases, as shown with GSVA. Altogether, these results point to multi-factorial changes in AD astrocytes, encompassing changes in astrocyte-neuron interactions and organelle dysfunction (model in **Fig. 7A** and description in **Discussion**).

**Figure 7.**
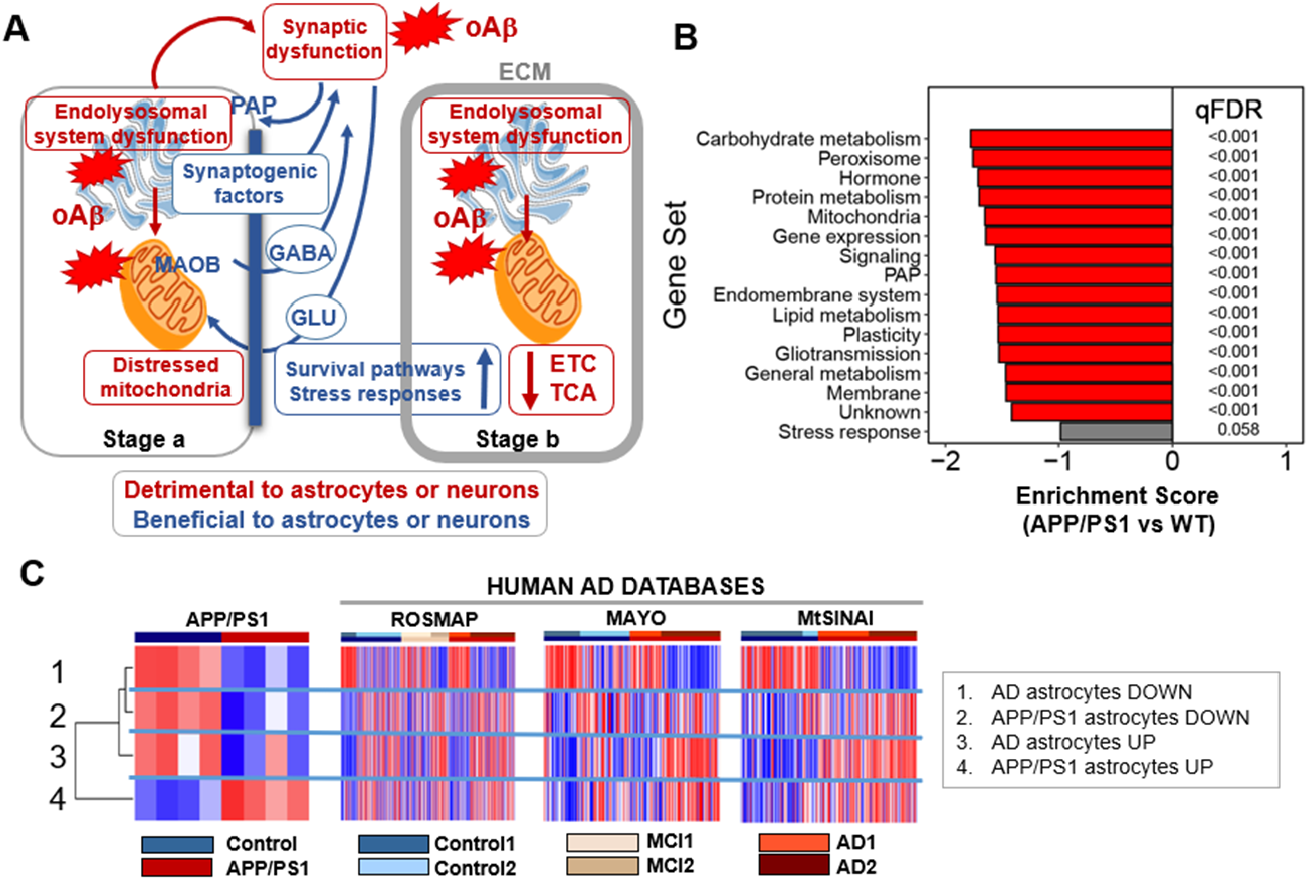
Model of cortical human AD astrocytes and comparison with mice. **A**. The model revolves around two predictions. *First prediction*: amyloid-β drives the phenotypical change of cortical astrocytes in AD by direct and indirect mechanisms. <u>Direct</u>: Amyloid-β causes endolysosomal and mitochondrial dysfunction in astrocytes, resulting in an adaptive astroprotective shift of gliotransmission to a GABA-predominant mode as a strategy to preserve mitochondrial respiration. <u>Indirect</u>: Synaptic dysfunction caused by amyloid-β in neurons augments the production of the PAP machinery and synaptogenic factors in astrocytes-a phenomenon of enhanced astrocytic plasticity to protect neurons. *Second prediction*: the phenotypical transformation of astrocytes in AD happens in early and late stages. Control2 subjects, cognitively normal, would be at stage a, and AD2 patients, diagnosed with dementia, at stage b. In stage a, the astroprotective change in glutamate/GABA fluxes may preserve neural-circuit homeostasis, because the GABA produced by MAO-B may locally compensate for the deficits in inhibitory tone that disrupts circuitry synchronization (Lee et al., 2020). By contrast, deficits in the endolysosomal system would result in partial phagocytosis of amyloid-β (Sollvander et al., 2016) and dystrophic neurites (Gomez-Arboledas et al., 2018), thus exacerbating amyloid-β accumulation, and hindering neuronal repair due to lack of debris elimination. In stage b, the mechanisms triggered to protect mitochondria may fail, and cortical astrocytes become deeply malfunctional, as suggested by the reduction in TCA cycle and ETC transcripts, despite the increase in protective stress responses and pro-survival pathways aimed at preventing astrocyte demise. For simplicity, only predominantly altered functions are depicted. ECM, extracellular matrix; ETC: electron transport chain, GLU: glutamate; oAβ: oligomeric amyloid-β; PAP: perisynaptic astrocyte processes; TCA: tricarboxylic acid. Figure adapted from Servier Medical Art (https://smart.servier.com/). **B**. GSVA of the transcriptome of astrocytes isolated from APP/PS1 mouse cortices using our manually curated functional categorization shows downregulation of astrocyte-enriched functions. **C**. Left, GSVA of transcriptome of APP/PS1 astrocytes according to the top dysregulated astrocyte genes in PC1 from the ROSMAP database (**Fig. 6B**). Astrocyte-enriched genes dysregulated in AD (up and down) are down-regulated in APP/PS1 astrocytes, as shown in B. Right, GSVA of PC1 AD and mouse APP/PS1 DEG gene sets in ROSMAP, MAYO and MtSINAI whole-brain transcriptomes shows that both the up and down-regulated mouse genes pools tend to be up-regulated in AD databases.

### Comparison with other human studies

Comparisons among our study and snRNAseq and whole-tissue transcriptomic studies (Mathys et al., 2019; B. Zhang et al., 2013) (Grubman et al., 2019) are in (**Supplementary file 11**). The greatest concordance was found between our study and (B. Zhang et al., 2013). Thus, there was an 11% overlap between cluster # 4 and Zhang data. Importantly, 94.9 % of the common genes were localized in the UP portion of the PC1 list, representing 28% of such list, and they were particularly enriched in the 50 Top UP PC1 (50% overlap with Zhang’s list). Common genes were related to ‘Gliotransmission’ (e.g., glutamate transporter *SLC1A3*), ‘Plasticity’ (e.g., Hippo Signaling *TP53BP2*), ‘Stress response ‘(e.g., antioxidant *PON2*), and ‘Mitochondria’ (e.g., amino acid metabolism *HADHB*). By contrast, there was scarce representation of astrocytic genes from (B. Zhang et al., 2013) in the down-regulated portion of our cluster 4, where the majority of the genes related to mitochondrial metabolism and the endolysosomal system lie. Since the Zhang astrocytic cluster was generated by co-expression within AD transcriptomes, we reason that down-regulated astrocytic genes may not co-express any longer with up-regulated ones. In summary, our study confirms the relevance of glutamate and amino acid metabolism in AD pathogenesis reported in large-scale network analysis (B. Zhang et al., 2013), and links it to organelle dysfunction, particularly that of mitochondria, which, as noted, presented a marked negative correlation with pathological stages according to the regressions with Braak/CERAD scores (p-value 0.0000203/0.000742, **Supplementary file 8**). As to scRNASeq studies, 12.7% and 1.1% of Cluster # 4 overlap with astrocytic DEG (Grubman et al., 2019; Mathys et al., 2019), respectively (**Supplementary file 11**). Comparison of the top 50 UP and 50 DOWN PC1 genes reveals little concordance: as compared to the 50% of top 50 PC1 genes found in (B. Zhang et al., 2013), only 0.3% and 1.3% of top 50 PC1 genes are annotated in the DEG in (Mathys et al., 2019) and (Grubman et al., 2019). Of note, (Mathys et al., 2019) and (Grubman et al., 2019) only share 32 genes, representing 47% of the DEG in (Mathys et al., 2019) and 4.5 % of those in (Grubman et al., 2019). Further, only 2.3% and 14.6% of the astrocytic cluster from (B. Zhang et al., 2013) overlap with (Mathys et al., 2019) and (Grubman et al., 2019). The incomplete equivalence among databases may be due to several factors, including: (i) differences in methodologies, (ii) classifications of disease (e.g., clinical in (B. Zhang et al., 2013), (Grubman et al., 2019) and our study, but associated to amyloid-β pathology in (Mathys et al., 2019) regardless of clinical diagnosis), (iii) brain regions sampled (prefrontal or temporal cortex in (Mathys et al., 2019) (B. Zhang et al., 2013) and in our case, but entorhinal cortex in (Grubman et al., 2019)), and/or (iv) the total number of subjects analyzed (1647 in (B. Zhang et al., 2013), 48 in (Mathys et al., 2019), 12 in (Grubman et al., 2019), and 766 herein.

### Cortical APP/PS1 astrocytes partially mimic cortical AD astrocytes

We finally asked whether changes identified in cortical AD astrocytes are recapitulated in mouse models. To answer this question, we examined the expression of human astrocytic genes from cluster #4 in the transcriptome of cortical astrocytes isolated from 15-18 month-old APP/PS1 versus WT mice (N=4 per group) (Orre et al., 2014), using the same functional categorization and statistical tools utilized for human AD astrocytes. Comparison with recent snRNAseq mouse data (Habib et al., 2020), is not appropriate since this study was carried in hippocampi. We found robust down-regulation of all human astrocytic functions in APP/PS1 cortical astrocytes (**Fig. 7B**). Thus, APP/PS1 astrocytes mimic the down-regulation in ‘endomembrane system’ and ‘mitochondria’ detected in AD astrocytes, but not the up-regulation of ‘PAP’, ‘stress responses’ and ‘plasticity’. We also compared the top 50 and bottom 50 DEG from APP/PS1 astrocytes (Orre et al., 2014) with the top 50 and bottom 50 genes from PC1 in human AD astrocytes (**Fig. 6B**). GSVA confirmed that both the top and bottom human AD astrocyte gene sets were down-regulated in APP/PS1 mice (**Fig. 7C**, left). Importantly, the pool of immune-related genes reported to be up-regulated in mouse APP/PS1 astrocytes (Orre et al., 2014) are predominantly microglial according to τ scores, and, with the exception of *C1S*, do not coincide with functionally related human astrocyte-enriched genes in cluster # 4 subcategories such as ‘cytokines’ and ‘complement’ (**Supplementary file 12**). As also concluded in (Orre et al., 2014), this divergence points to the existence of two distinct gene pools in cortical APP/PS1 astrocytes that change in opposite directions in advance disease stages: a down-regulated pool of typical homeostatic astrocyte genes, and an up-regulated pool of typical microglia genes. The loss of astrocytic functions and the adoption of a microglia-like phenotype suggests that cortical APP/PS1 astrocytes may undergo a phenotypical involution with time.

Finally, regardless of the direction of change in mice, DEG in APP/PS1 vs WT cortical astrocytes were largely up-regulated in AD2 the three human databases (**Fig. 7C**, right panels). This suggests predominance of typical microglial genes in advanced AD, plausibly upregulated in both microglia and astrocytes.

## Discussion

Because of the challenges associated with the isolation of astrocytes or their nuclei, and to profit from the wealth of large publicly available human datasets, the goal of this study was to gain insight into what happens to astrocytes in AD using whole-brain transcriptomes and systems-biology tools. The approach consisted of re-compartmentalizing whole-brain transcriptomes from Control, MCI and AD subjects into contributions from individual cell types by using pre-clustered cell-specific gene sets. The study has four main findings. First, GSVA and PCA show that AD astrocytes—so defined because they appear in patients with the typical down-regulation of neuronal synaptic functions documented in AD—undergo a profound phenotypical transformation encompassing down-regulation of genes encoding for organelle proteins, and up-regulation of genes related to PAP, gliotransmission, plasticity and stress responses. Second, unbiased hierarchical clustering of patient cohorts reveals major intra-cohort heterogeneity, i.e., relationships are not robust between gene profiles, clinical diagnosis, and pathological stage according to CERAD and Braak scoring, as recently reported in (Neff, 2021). In other words, some non-demented subjects present AD-like astrocytes and neurons, while some demented subjects present healthy ones, particularly in MAYO and ROSMAP. Third, according to regression analyses between functions and CERAD or Braak scores, some astrocytic functions may be altered early in the AD continuum (e.g., PAP, gliotransmission and endomembrane system), while other functions may be increasingly dysregulated in parallel with the deposition of neuritic plaques and neurofibrillary tangles (e.g., mitochondrial functions, plasticity-related pathways and stress responses). Fourth, multivariate statistics and hierarchical clustering show that cortical astrocytes from human AD and from a 15–18-month-old mouse model of ADAD (Orre et al., 2014) are distinct transcriptional entities.

### Limitations

A limitation of our study is that pathway dysregulation during AD may result in the remodeling of cell-type specific gene clusters, such that gene clusters identified in healthy brain transcriptomes may no longer exist in AD. However, quantification of network reorganization in AD subjects using a metric called modular differential connectivity (MDC) showed equal or enhanced connectivity in 95.5% of the modules (B. Zhang et al., 2013), suggesting that clusters identified by co-expression analyses in healthy cells are, for the most part, preserved in AD. Nevertheless, we recognize that our analysis may overlook changes in transcripts ubiquitous in all cell types, e.g., glycolysis related transcripts. It may also overlook upregulation in astrocytes of transcripts typical of non-astrocytic cells, namely microglia, including the inflammation-like response common to astrocytes and microglia, as described in 15-18-month-old APP/PS1 transgenic mice (Orre et al., 2014). Conversely, our analysis may detect down-regulated astrocytic genes (e.g., mitochondrial genes) that may go unnoticed in large-scale clustering, for their expression changed in opposite directions to the majority of genes in the original cluster (B. Zhang et al., 2013). Still, our results may be just the tip of the iceberg in terms of defining the totality of changes in human AD astrocytes.

Yet another limitation of the analysis is that it does not reveal astrocyte populations, recently described in a mouse model of AD using snRNAseq (Habib et al., 2020), although it is tempting to speculate that the four astrocytic gene clusters identified in the clinical cohorts (**Fig. 3**) might represent distinct transcriptomic states. Further, because transcriptomic data are cross-sectional and descriptive, they do not demonstrate cause-effect relationships between pathway dysregulation and pathology, or unequivocally prove that changes are adaptive (meant to compensate for dysregulated functions and maintain or restore homeostasis), maladaptive (irreversibly contributing to dyshomeostasis and neurodegeneration), or epiphenomena with scarce bearing on disease onset and progression. Finally, genetic data does not directly inform about changes in proteins and metabolites. Hence, predictions, based upon these data (below) need validation in appropriate models.

### Key implications and predictions

#### Use of molecular data for subject stratification in clinical trials

Neurocognitive measurements, atrophy patterns assessed with neuroimaging, neurofibrillary tangles assessed *postmortem*, CSF levels of amyloid-β1-42 and Tau, as well as Tau neuroimaging, have revealed subtypes of AD (Devi & Scheltens, 2018; Dujardin et al., 2020). This clinical and biological heterogeneity, which is increasingly recognized to be a major obstacle to establishing drug efficacy in clinical trials, starts to be addressed with molecular tools, as shown by the recent detection of three molecular classes in an extended MtSINAI cohort using RNAseq transcriptomics in four brain regions (Neff, 2021). They found the largest global change in the parahippocampal gyrus (PHG), presenting three molecular subtypes among subjects, termed A, B and C. Although our analysis was performed in cortical samples rather than the PHG, we reason that our AD2 group might correspond to what they called ‘C typical’ AD class, and our AD1 might correspond to what they called the ‘A atypical’ AD class. The reason is that both AD2 and C present downregulation of synaptic networks as compared to AD1 and A. No further comparison between our study and (Neff, 2021) can be performed with regards to astrocytes because, among only three genes reported to be upregulated in astrocytes in the ‘C typical AD class’, only *GNA12* appears in our astrocytic cluster # 4—although it is indeed upregulated in ROSMAP (**Supplementary data 13**).

A disconnect between presence of AD pathology and cognition has been long recognized (Neuropathology Group. Medical Research Council Cognitive & Aging, 2001). The terms ‘resistance’ and ‘resilience’ have been coined to describe two distinct clinical scenarios consisting of avoiding pathology (’resistance’) *versus* coping with pathology (’resilience’) (reviewed in (Arenaza-Urquijo & Vemuri, 2018)). Thus, resistant subjects would be cognitively normal without significant Abeta or Tau pathology despite being at high-risk, considering several factors including age or APOE genotype. By contrast, resilient subjects would remain cognitively normal despite significant AD pathology. We speculate that our AD1 subjects were perhaps ‘resilient’ to AD pathology, which they had, according to high CERAD and Braak scores, but that they developed dementia due to comorbidities (e.g., Lewy bodies, vascular disease, TDP43 deposits). However, the possibility exists that the tissues used for transcriptomic analysis and neuropathological assessment of neuritic plaques and neurofibrillary tangles were not the same. This means that the transcriptomic analysis in demented AD1 subjects with a healthy-like molecular profile might have been performed in cortical tissue spared from ongoing pathology, such that AD1 subjects were not truly resilient. By contrast, regardless of how omics and neuropathological assessments were performed, Control2 subjects in ROSMAP and MAYO with an AD-like molecular profiles were cognitively normal. Thus, Control2 subjects might be high-risk, pre-symptomatic, resistant subjects in whom the appearance of neuritic plaques, Tau tangles, and cognitive impairment, was delayed despite the decreased expression of synaptic genes, plausibly caused by soluble oligomeric amyloid-β (Marsh & Alifragis, 2018). Whatever the case, our study supports the use of omics-based molecular phenotyping and clustering statistics as unbiased tools to stratify patients in clinical trials in the spirit of personalized medicine.

#### Early dysregulation of the endolysosomal system in astrocytes

The category *‘*endomembrane system’ was found down-regulated in AD2 vs Control1 by GSVA. Further, genes related to endoplasmic reticulum (e.g., *FAF2, PIGX*), endocytosis (e.g., *ATXN2, MEGF8, RAB11FIP2*), and autophagy (*ATG9A*) ranked among the top50PC1 downregulated in ROSMAP (**Fig. 6B**), and were significantly down-regulated in at least two of the three databases by Wilcoxon test (**Supplementary file 4**). These findings support cumulative evidence in mouse and *in vitro* models of AD pointing to inefficient phagocytosis and degradation of amyloid-β protofibrils (Funato et al., 1998; Sanchez-Mico et al., 2020; Sollvander et al., 2016) and dystrophic neurites (Gomez-Arboledas et al., 2018) by astrocytes, as well as decreased exosome production from astrocytes (Abdullah et al., 2016). Thus, our study and others do not support the notion that astrocytes efficiently clear amyloid-β plaques, as initially suggested by evidence in non-primary cultured astrocytes (Wyss-Coray et al., 2003), which, arguably, are endowed with greater motility and morphological plasticity than primary or *in situ* astrocytes. Moreover, targeted enhancement of lysosomal biogenesis by overexpression of the transcription factor EB (TFEB) in astrocytes enhances Tau clearance, and reduces pathological hallmarks in the hippocampus of PS19 tauopathy mice (Martini-Stoica et al., 2018), suggesting that the improvement of lysosomal function in astrocytes may also reverse Tau pathology in AD. Along these lines, TFEB has been reported to regulate AD risk genes in an astrocyte subpopulation detected in AD brains by snRNAseq (Grubman et al., 2019).

#### Early and concomitant alteration of neuronal-astrocyte contacts

This prediction is based on two findings: the inverse correlation of changes in synaptic functions and PAP, and the lack of correlation between these functional categories with pathological stages, suggesting that they change early in AD. Accordingly, both PAP and synaptic genes are dysregulated in non-demented subjects with a *type 2* molecular profile (Control2), which, as noted earlier, may represent preclinical AD. Specifically, GSVA showed up-regulation of ‘PAP morphology’, whose leading gene *EZR* encodes ezrin, a protein involved in PAP motility and in the anchoring of the astrocyte membrane to neurons or the extracellular matrix (Derouiche & Geiger, 2019). Also, genes related to integrin and cell motility (e.g., *FERMT*2, *PREX2*) were part of the top genes detected by PCA as defining AD astrocytes (**Fig. 6B**). We posit that upregulation of PAP genes is an adaptive change to improve the coverage of neurons to counteract the down-regulation of synaptic elements directly caused by oligomeric amyloid-β (Marsh & Alifragis, 2018). This scenario is supported by the upregulation of genes encoding for synaptogenic factors such as thrombospondin and glypicans (*THSD1, GPC4*) in the three databases (**Supplementary file 4**).

#### Mitochondrion dysfunction in AD astrocytes may impact gliotransmission

It is generally assumed that the well-established mitochondrion dysfunction (Swerdlow, 2018) and reduced glucose metabolism in AD (often referred to as ‘hypometabolism’ (Hoffman et al., 2000)) are neuronal phenomena. However, our analysis reveals down-regulation of nuclear-encoded mitochondrial genes encoding TCA cycle and ETC components in astrocytes, suggesting deficiencies in energy-generating mitochondrial pathways. For example, one of the consensus downregulated genes, *ATPAF1*, is a component of the ATP synthase (complex V), and it is known that impairment of the ETC beyond complexes I-III increases reactive-oxygen species formation while decreasing ATP formation (Shi & Gibson, 2007). Moreover, i*n vitro* and *ex vivo* studies link mitochondrial dysfunction in astrocytes to amyloid-β. Thus, in co-cultures of rat hippocampal neurons and astrocytes, amyloid-β causes loss of mitochondrial potential and transient mitochondrial depolarization, concomitant to activation of NADPH oxidase and decrease glutathione production, in astrocytes, but not neurons (Abramov, Canevari, & Duchen, 2004). Loss of membrane potential and depolarization of astrocytic mitochondria is prevented by antioxidants, and by the use of glutamate by mitochondrial complex I as a substrate, suggesting that amyloid-β-elicited mitochondrial dysfunction is caused by oxidative stress and deficits in substrate supply (Abramov et al., 2004). Interestingly, the mitochondrial glutamate transporter *SLC25A18*, as well as the plasma membrane glutamate transporters *SLC1A3* and *SLC7A11* are up-regulated in the three databases, and *SLC6A11* in two (**Supplementary file 4**), suggesting that AD astrocytes recycle glutamate to counteract the damage caused by amyloid-β to astrocytic respiration by increasing the supply of fuels amenable for oxidation. Along these lines, *HADH8*, which encodes for an enzyme involved in mitochondrial beta oxidation, was found upregulated in AD astrocytes by PCA.

Likewise, the consensus upregulation in AD astrocytes of the monoamine oxidase *MAO-B*, a dopamine-degrading enzyme located in the outer mitochondrial membrane, may be interpreted as an attempt to reverse respiratory deficits. This is supported by the recent discovery that MAO-B increases the polarization of the inner mitochondrial membrane and ATP production by shuttling electrons through the inter-membrane space (Graves et al., 2020). MAO-B upregulation in AD is relevant for two reasons. First, the MAO-B ligand (11)C-deuterium-L-deprenyl may serve as a PET-based biomarker of early astrocyte dysfunction in AD, as documented in ADAD (Rodriguez-Vieitez et al., 2016). Second, MAO-B may affect the excitatory/inhibitory balance of neural circuits by producing GABA from putrescine, as shown in APP/PS1 mice (Jo et al., 2014). In support of this scenario, the genes encoding the GABA transporter *SLC6A11/GAT3*, which may extrude GABA coupled to glutamate uptake, and *SLC7A2*, which imports the putrescine precursor L-arginine, are consensus dysregulated genes (**Supplementary file 4)**. Taken together, the data support that impaired mitochondrial respiration in astrocytes switches gliotransmission to a GABA-dominant mode. A detrimental effect of astrocytic GABA has been proposed based on the beneficial effects of the MAO-B inhibitor selegiline on electrophysiological readouts in APP/PS1 mouse hippocampi (Jo et al., 2014); however, there is no benefit of selegiline in patients (Birks & Flicker, 2003), thus raising doubts as to whether APP/PS1 mice appropriately model the impact of astrocytic GABA in AD. Alternatively, astrocytic GABA might counteract the loss of GABAergic tone that has been causally related with neuronal hyperactivity, desynchronization of neural circuits, and amyloid-β production (Lee, Gerashchenko, Timofeev, Bacskai, & Kastanenka, 2020). Since our regression analyses reveal that mitochondrial dysregulation in astrocytes progressively increases in advanced pathological stages, and since the upregulation of MAO-B detected by PET in the prodromal stages in ADAD is transient (Rodriguez-Vieitez et al., 2016), we reason that the tw0 strategies used by mitochondria to preserve membrane potential and ETC (i.e., enhanced supply of substrates such as aromatic amino acids like glutamate, and of electrons *via* MAO-B) eventually fail in astrocytes. As a consequence, glutathione production might be decreased and NADPH oxidase activity increased (Shi & Gibson, 2007), perhaps exacerbating oxidative stress and overall astrocyte malfunction in advanced AD.

#### Pathological-stage dependent upregulation of ‘plasticity’ and ‘stress response’ pathways might explain why, unlike neurons, astrocytes survive in AD

The progressive upregulation of the subcategory ‘ECM’, which includes integrins and proteoglycans, is not surprising considering the indisputable adoption of a ‘reactive’ morphology by astrocytes in AD, consisting of engrossment of primary and secondary processes due to over-expression of the intermediate filament protein GFAP, and increased production of ECM proteins (Escartin, Guillemaud, & Carrillo-de Sauvage, 2019). More intriguing is the consensus up-regulation of hippo signaling, an evolutionarily conserved network with a central role in the regulation of cell proliferation, cell fate, organ growth and regeneration (Misra & Irvine, 2018), and no reported role in healthy adult astrocytes, despite the fact that some of the genes in this pathway are highly specific to human adult astrocytes according to τ (e.g., *WWC1* τ =0.91; *WWOX* τ =0.86, and *YAP1* τ =0.88). *YAP1* was, moreover, detected by PCA as a relevant gene in the astrocytic phenotype in AD. The upregulation of hippo signaling in AD astrocytes, as well as the global upregulation of genes involved in brain development may be a manifestation of the well documented phenomenon of re-activation of developmental pathways in reactive astrocytes that some authors interpret as a failed attempt at reprogramming into neural stem cells (Torper & Gotz, 2017), and we interpret to be a protective response to facilitate the survival of defective astrocytes. In addition, the recent observation that *YAP1* regulates scar-border formation in spinal cord injury in mice (Xie et al., 2020), suggests that hippo signaling is coordinated with the increased production of ECM in AD astrocytes. Finally, the consensus dysregulation of the subcategory ‘TNF-alpha’ (**Supplementary file 10**), and the consensus upregulation of *TNFRSF11B, TNFSF13 and TRAF3IP2*, members of the IL17 family (*Il17D, IL17RB, and IL17RD*), and complement factors (*CD59, C1S*) (**Supplementary file 4**) might be interpreted as detrimental ‘neuroinflammation’, but evidence supports modulatory or protective actions of these factors (Masgrau, Guaza, Ransohoff, & Galea, 2017). First, TNFα may facilitate the release of glutamate by improving vesicle docking at the astrocyte plasma membrane prior to exocytosis (Santello et al., 2011), such that TNFα signaling may help adjust gliotransmission in AD astrocytes. Second, the finding that Nrf2/IL17D axis acts as an antioxidant protective pathway in stress responses induced by tumorigenic stimuli and viral infections (Saddawi-Konefka et al., 2016), supports an astroprotective role of members of the IL17 family in AD. Also, members of the complement system have been shown to facilitate amyloid-β phagocytosis (Iram et al., 2016).

### Model of the phenotypical transformation of cortical astrocytes in AD

The key predictions from our analysis are summarized in **Fig. 7A**. We posit that amyloid-β drives the phenotypical change of astrocytes in AD in several stages by causing dysfunction of the endolysosomal/mitochondrial axis, which, in turn, prompts a change in the balance of excitatory/inhibitory neurotransmission. Impairment of mitophagy due to endolysosomal malfunction may further exacerbate mitochondrial dysregulation. Control2 subjects (cognitively normal/AD-like molecular profile) would be at stage a, and AD2 patients (demented, AD molecular profile) at stage b. In both a and b, the phenotypical change is complex and *mixed* with regards to possible clinical impact, for changes that may disrupt homeostasis of astrocytes themselves and/or astrocyte-neuron communications coexist with changes that may protect astrocytes and/or their interactions with neurons. The model thus represents a departure from simplistic neuroprotective/neurotoxic classifications of reactive astrocytes from the point of view of neurons. As recently discussed (Escartin et al., 2021), an ‘astrocyte-centric’ stance focused on the clarification of the maladaptive or adaptive nature of pathway changes in the complex astrocyte biology should be adopted. What is then the *net* impact of the mixed functional changes of reactive astrocytes on AD? In short, we posit that, despite organelle dysfunction, astrocytes manage in early disease stages to partially preserve their functions, including the modulation of neural circuits, through adaptive changes, while they become globally malfunctional in advanced stages. Thus, the early changes in astrocyte-neuron interplay may delay the onset of clinical symptoms, while detrimental phenomena would prevail in stage b. If these predictions are correct, therapies aimed at protecting and restoring the functions of the endolysosomal system and mitochondria to halter the transformation of astrocytes to stage b might help arrest the progression of AD. Even if GABA were detrimental, as reported in (Jo et al., 2014), preservation of mitochondrial respiration is therapeutically indicated, because increased GABA production, whatever its effect, would be the result of mitochondrial impairment. In summary, the present study points to prevention of organelle dysfunction in astrocytes as a therapeutic strategy in AD. These predictions need to be tested in transgenic mice or human cellular models that appropriately recapitulate the complex transformation of astrocytes in the AD continuum.

## Conclusions

Our novel systems-biology based identification of astrocytic clusters in three independent datasets, encompassing 766 patients, represents the most comprehensive transcriptomic analysis of human astrocytes in AD to date. We highlight that manually curated functional annotations were implemented to circumvent deficits in astrocyte-specific annotations in current platforms. Further, molecular heterogeneity of subjects with the same clinical diagnosis was detected by hierarchical clustering and controlled for by performing independent statistics. The findings have led to a model of stage-dependent astrocyte dysfunction in AD caused by Aβ-elicited damage of the endolysosomal and mitochondrial systems. If this model is correct, astrocyte-targeted therapies aimed to prevent organelle malfunction in astrocytes may be beneficial for AD.

## Supporting information

Suppplementary file 1

Suppplementary file 2

Suppplementary file 3

Suppplementary file 4

Suppplementary file 5

Suppplementary file 6

Suppplementary file 7

Suppplementary file 8

Suppplementary file 9

Suppplementary file 10

Suppplementary file 11

Suppplementary file 12

## Data Availability

All datasets are public and identifiers are noted for each dataset used.

## Acknowledgements

The authors thank Dr. Alberto Lleó of the Unitat de Memòria at the Hospital Sant Pau, Barcelona, Spain, for critical reading of the manuscript. This work was supported by Fundació Marato-TV3 (grant number 201414-30), and Grup de Recerca en demèncias: Sant Pau SGR2017547, Generalitat de Catalunya (E.G.), by funds from the Coins for Alzheimer’s Research Trust (L.B.W.) and startup funds from the Woodruff School of Mechanical Engineering at Georgia Tech (L.B.W.). L.D.W. and A.F.P were supported in part by the National Institutes of Health Cell and Tissue Engineering Biotechnology Training Grant (T32-GM008433). CNRS, CEA and France Alzheimer (C.E.). The results published here are in whole or in part based on data obtained from the AD Knowledge Portal (https://adknowledgeportal.synapse.org). Study data were provided by the following sources: The Mayo Clinic Alzheimers Disease Genetic Studies, led by Dr. Nilufer Ertekin-Taner and Dr. Steven G. Younkin, Mayo Clinic, Jacksonville, FL using samples from the Mayo Clinic Study of Aging, the Mayo Clinic Alzheimers Disease Research Center, and the Mayo Clinic Brain Bank. Data collection was supported through funding by NIA grants P50 AG016574, R01 AG032990, U01 AG046139, R01 AG018023, U01 AG006576, U01 AG006786, R01 AG025711, R01 AG017216, R01 AG003949, NINDS grant R01 NS080820, CurePSP Foundation, and support from Mayo Foundation. Study data includes samples collected through the Sun Health Research Institute Brain and Body Donation Program of Sun City, Arizona. The Brain and Body Donation Program is supported by the National Institute of Neurological Disorders and Stroke (U24 NS072026 National Brain and Tissue Resource for Parkinson’s Disease and Related Disorders), the National Institute on Aging (P30 AG19610 Arizona Alzheimer’s Disease Core Center), the Arizona Department of Health Services (contract 211002, Arizona Alzheimer’s Research Center), the Arizona Biomedical Research Commission (contracts 4001, 0011, 05-901 and 1001 to the Arizona Parkinson’s Disease Consortium) and the Michael J. Fox Foundation for Parkinson’s Research. Study data were also provided by the Rush Alzheimer’s Disease Center, Rush University Medical Center, Chicago. Data collection was supported through funding by NIA grants P30AG10161 (ROS), R01AG15819 (ROSMAP; genomics and RNAseq), R01AG17917 (MAP), R01AG30146, R01AG36042 (5hC methylation, ATACseq), RC2AG036547 (H3K9Ac), R01AG36836 (RNAseq), R01AG48015 (monocyte RNAseq) RF1AG57473 (single nucleus RNAseq), U01AG32984 (genomic and whole exome sequencing), U01AG46152 (ROSMAP AMP-AD, targeted proteomics), U01AG46161(TMT proteomics), U01AG61356 (whole genome sequencing, targeted proteomics, ROSMAP AMP-AD), the Illinois Department of Public Health (ROSMAP), and the Translational Genomics Research Institute (genomic). ROSMAP data can be requested at www.radc.rush.edu. All data generated during this study are included in this published article and its supplementary information files. Codes are available upon reasonable request.

## Supplementary files

**Supplementary file 1. Generation of cell-specific gene modules**. Tabs:

1. Tab description.
2. Transcriptomic databases of human-brain cells obtained from (Y. Zhang et al., 2016).
3. PCA of transcriptomic data of astrocytes isolated from young (<47 years old) and old (>47 years old) shows that they were significantly different.
4. Specific data used for τ scoring and hierarchical clustering.
5. Top τ scores per gene, indicating which cell type presents that score; and pie charts of the distribution of specific and enriched genes among brain cells.
6. Raw results of hierarchical clustering.
7. Cluster description.
8. Cluster selection.

**Supplementary file 2. Functional categories of astrocytic and neuronal modules**. Tabs:

1. Circo plots showing functional categories (inner ring) and subcategories (outer ring) of astrocytic gene cluster 4 (1650 genes) and neuronal gene cluster 13 (1258 genes). For astrocytes, there are 17 groups and 133 subgroups. For neurons, 15 groups and 76 subgroups.
2. Functional categorization of neuronal cluster 13.
3. Functional categorization of astrocytic cluster 4.

**Supplementary file 3. Hierarchical clustering of astrocytic and neuronal gene sets before and after data normalization**. Gene expressions were normalized according to the probabilistic distribution of relative expressions to reach distribution peaks of zero (Methods). Normalization improved the resolution of astrocytic and neuronal gene clusters between and within cohorts.

**Supplementary file 4**. Cluster # 4 expression in the three databases after data normalization. The statistical comparison between AD2 and Control1 was performed by the Wilcoxon rank sum test. Statistical significance pFDR < 0.05. Consensus dysregulated genes are indicated in color: blue, down-regulated; yellow, up-regulated, and grey, unchanged.

**Supplementary file 5**. GSVA statistics of inter-cohort comparisons of changes in neuronal and astrocytic *general* functions in MAYO, ROSMAP and MtSINAI databases. Comparisons: ROSMAP: Control2 vs Control1 (pFDR1), MCI1 vs Control1 (pFDR2), MCI2 vs Control1 (pFDR3), AD1 vs Control1 (pFDR4), AD2 vs Control1 (pFDR4). MAYO and MtSINAI: Control2 vs Control1 (pBootFDR1), AD1 vs Control1 (pFDR2), AD2 vs Control1 (pFDR3). Tabs:

1. Functional categories in neurons.
2. Functional categories in astrocytes.

**Supplementary file 6. Extended Fig. 4B. Alteration of neuron-enriched functions in AD**. Enrichment scores of functional categories significantly changed in at least 2 out of 3 databases. (&) pFDR <0.01, (#) pFDR < 0.05 and (*) pFDR < 0.001, versus Control1.

**Supplementary file 7. Correlations between changes in neuronal general functional categories and pathological stages**. Bolded numbers are probability of a zero slope for linear model.

A. Braak scores.
B. CERAD scores.

**Supplementary file 8. Correlations between changes in astrocytic general functional categories and pathological stages**. Bolded numbers are probability of a zero slope for linear model.

A. Braak scores.
B. CERAD scores.

**Supplementary file 9. Correlations among altered functions in the ROSMAP, MAYO, and MtSINAI databases**.

**A, C, E**. Hierarchical clustering of Pearson correlation coefficients. The enrichment scores of all functions across all samples were correlated with one another (Methods). Clustering segregated correlations in two main branches that grouped down-regulated (branch 1) and up-regulated functions (branch 2), respectively. Functions found altered in at least two out of the three databases are labeled in blue (down-regulated) or red (up-regulated) squares. Strongest direct correlations were among highly related biological processes such as ‘membrane potential’, ‘neurotransmission’ and ‘synaptic plasticity’ in neurons, and ‘PAP’ and ‘gliotransmission’ in astrocytes (orange scale). The strongest anti-correlations (i.e., concomitant changes in opposite directions, grey scale) were: (i) down-regulation of synaptic functions versus up-regulation of neuronal ‘mitochondria’, ‘stress response’ and ‘gene expression’ (dark green squares); (ii) decrease in synaptic functions versus up-regulation of ‘PAP/gliotransmission’ and ‘plasticity’ in astrocytes (light green squares); (iii) in astrocytes, increases in ‘PAP/gliotransmission’ versus down-regulation of ‘endomembrane system’ and ‘gene expression’, and ‘plasticity’ versus ‘mitochondria’ (pink squares). In summary, anti-correlations (negative p-values) point to highly intertwined changes between morphological remodeling in astrocytes and neuronal dysfunction, and between survival responses and organelle dysfunctions in astrocytes.

**B, D, F**. Heatmap of FDR adjusted p-values. In red, adj FDR p-values < 0.05

**Supplementary file 10**. GSVA statistics of inter-cohort comparisons of changes in neuronal and astrocytic *subcategories* in MAYO, ROSMAP and MtSINAI databases. Comparisons: Control2 vs Control1 (pFDR1), AD1 vs Control1 (pFDR2), AD2 vs Control1 (pFDR3). Tabs:

1. All functional subcategories/MAYO.
2. All functional subcategories /ROSMAP.
3. All functional subcategories /MtSINAI.
4. PAP / all databases.
5. Gliotransmission / all databases.
6. Plasticity /all databases.
7. Stress response /all databases.
8. Mitochondria in all data /all databases.
9. Endolysosomal system /all databases.

**Supplementary file 11**. Astrocyte cluster # 4 ranked according to PC1 in ROSMAP, and comparison with DEG identified by snRNAseq in human AD astrocytes (Mathys et al., 2019) (Grubman et al., 2019). Tabs:

1. Venn diagrams.
2. Comparison of PC1/ROSMAP with (Mathys et al., 2019).
3. Comparison of PC1/ROSMAP with (Grubman et al., 2019).
4. Comparison of PC1/ROSMAP between (Mathys et al., 2019) and (Grubman et al., 2019).

**Supplementary file 12**. Comparison of the functional category of cluster # 4 of human astrocytes containing immunity-related genes with the immunity-related GO groups found significantly dysregulated in APP/PS1 mouse astrocytes (Orre et al., 2014). Only C1S (Complement Component 1, S Subcomponent) appears in both gene sets.

