## Supplementary figures and images for "Multi-transcriptomic analysis points to early organelle dysfunction in human astrocytes in Alzheimer’s disease"

### Suppplementary file 3

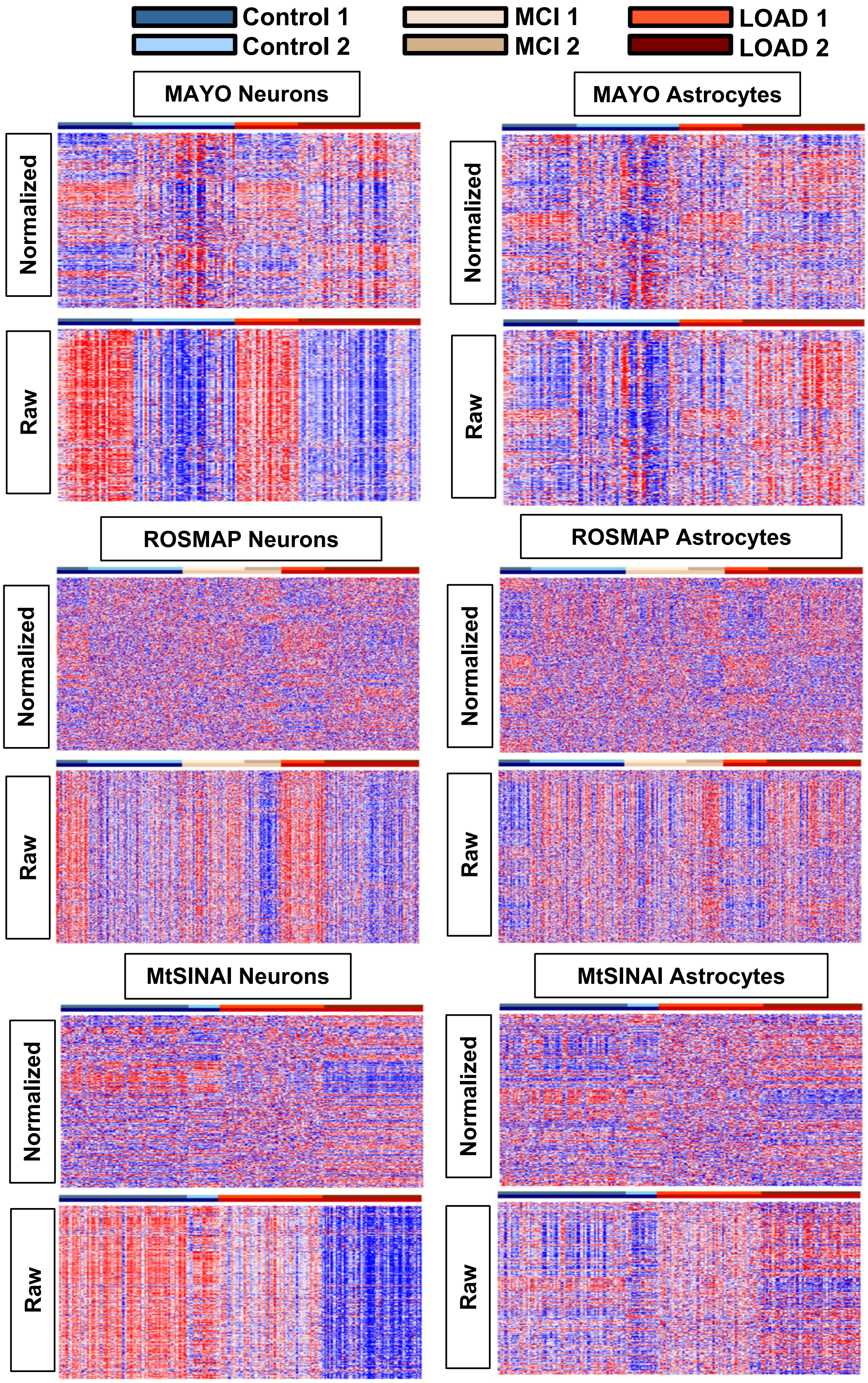


Supplemenary file 3

### Suppplementary file 6

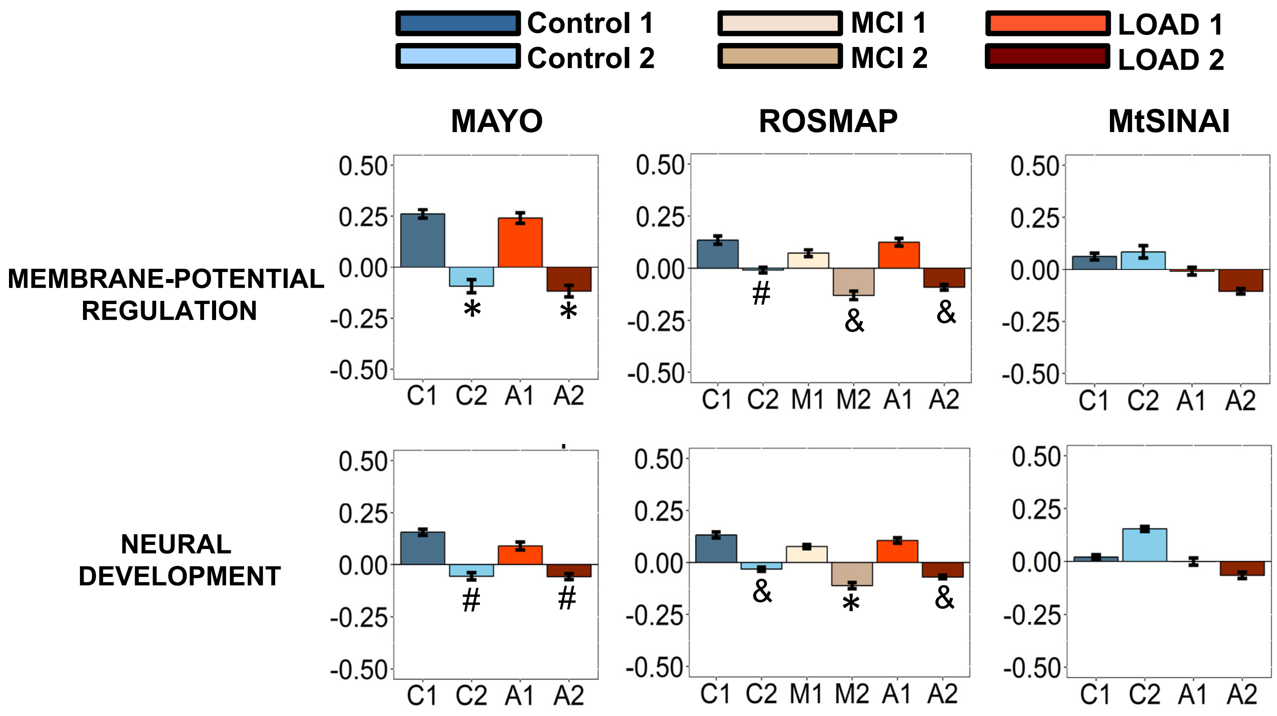


Supplementary file 6

### Suppplementary file 7

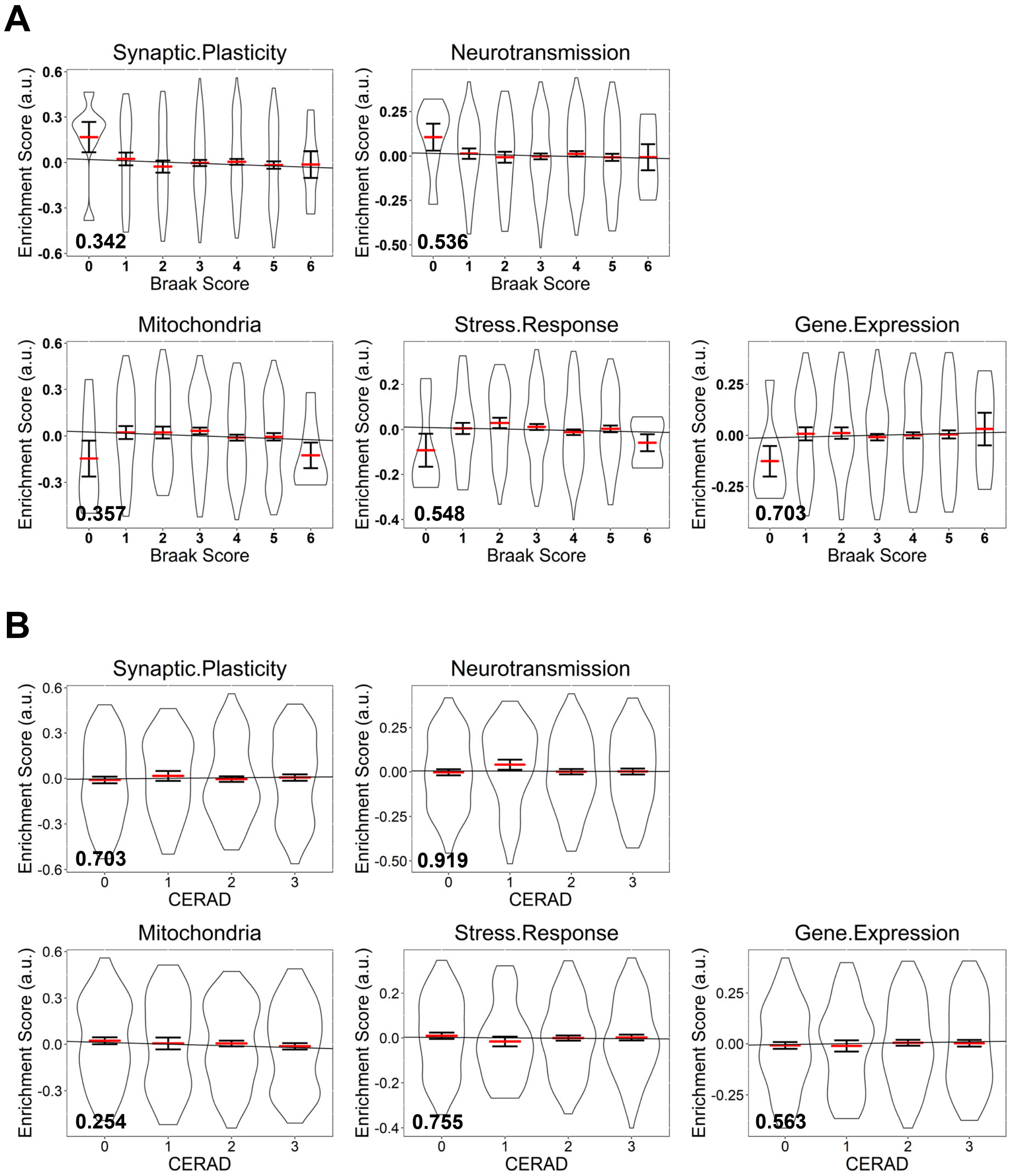


**Supplementary file 7**

### Suppplementary file 8

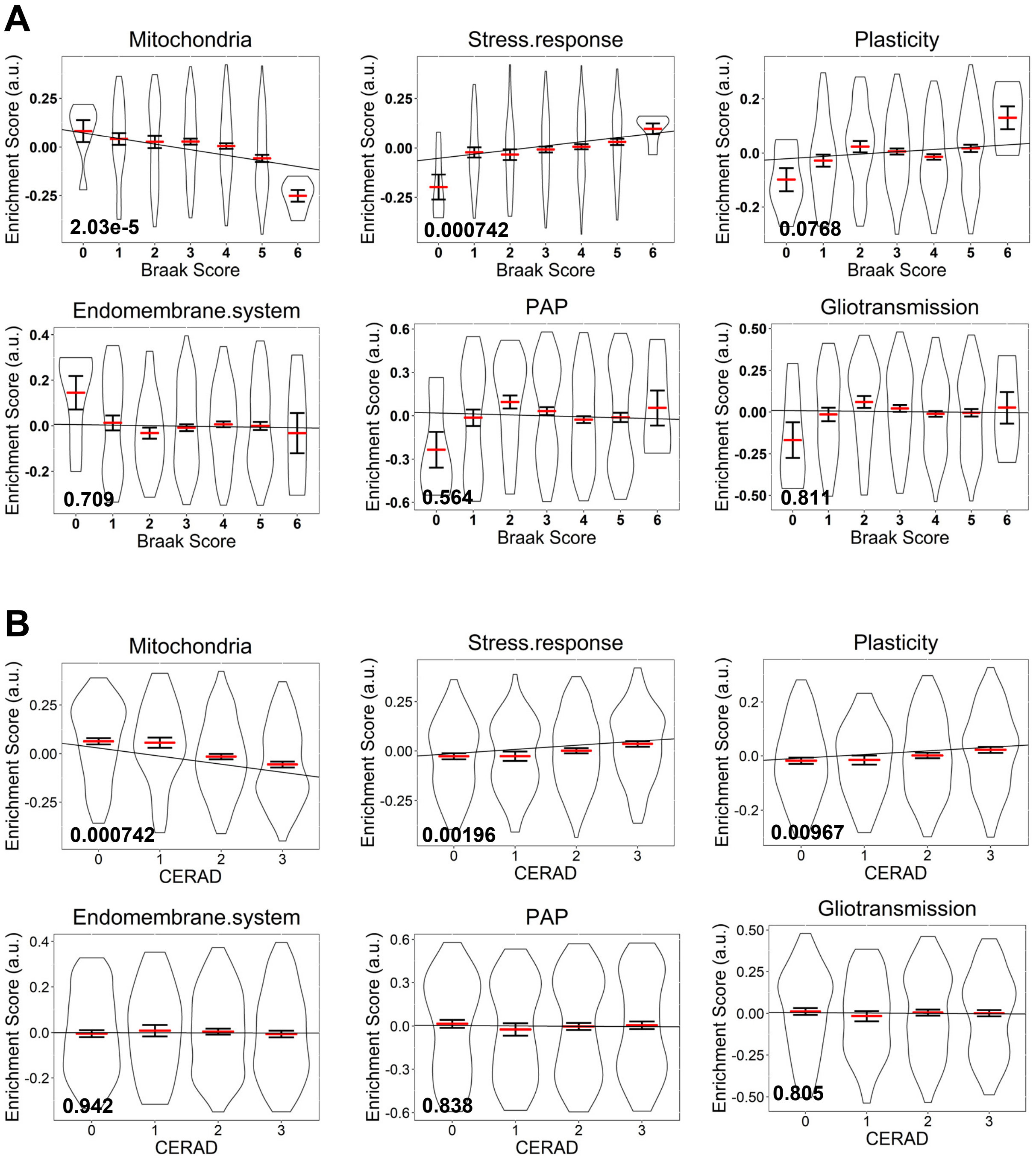


**Supplementary file 8**

### Suppplementary file 9

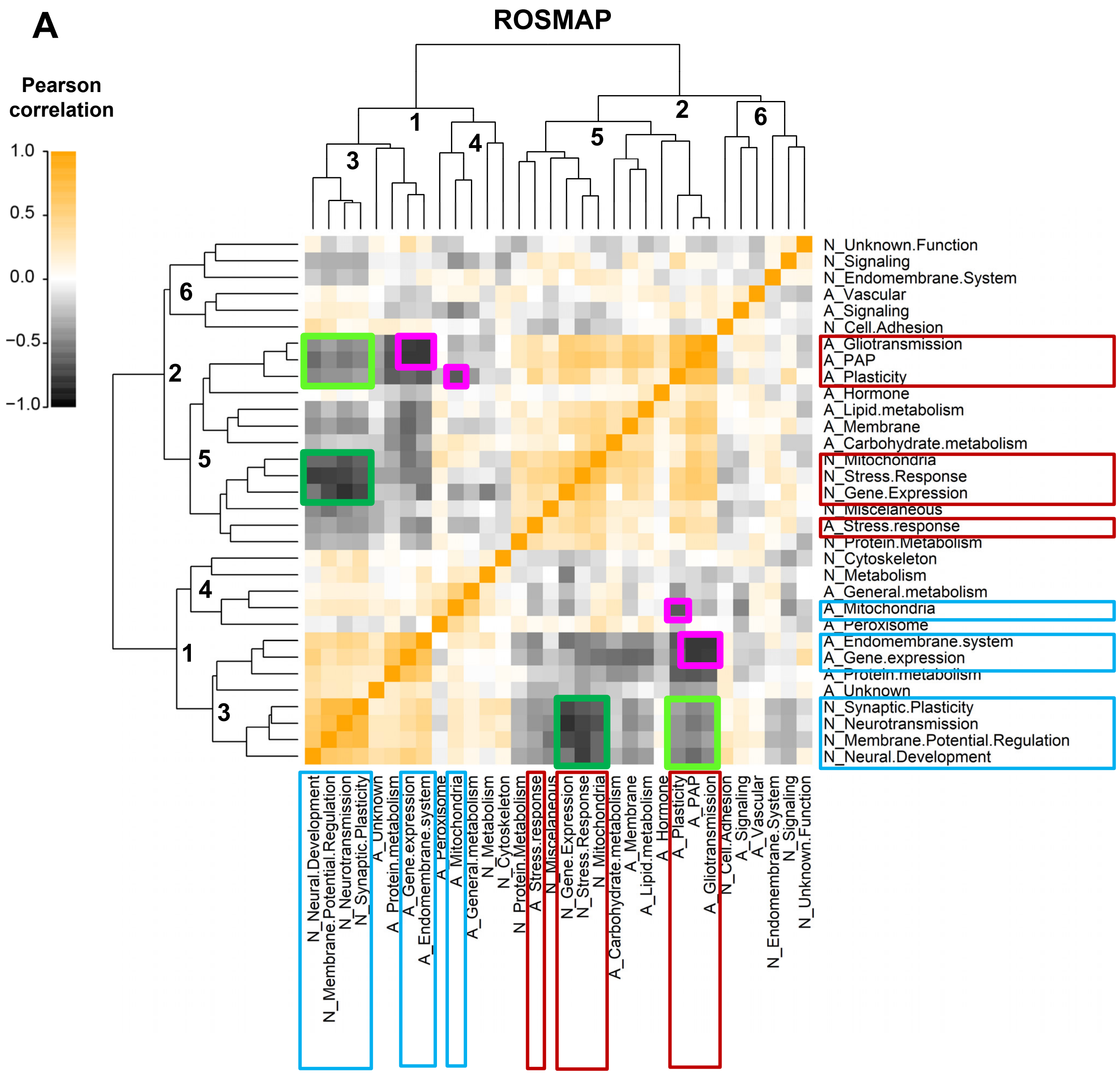


Supplementary file 9

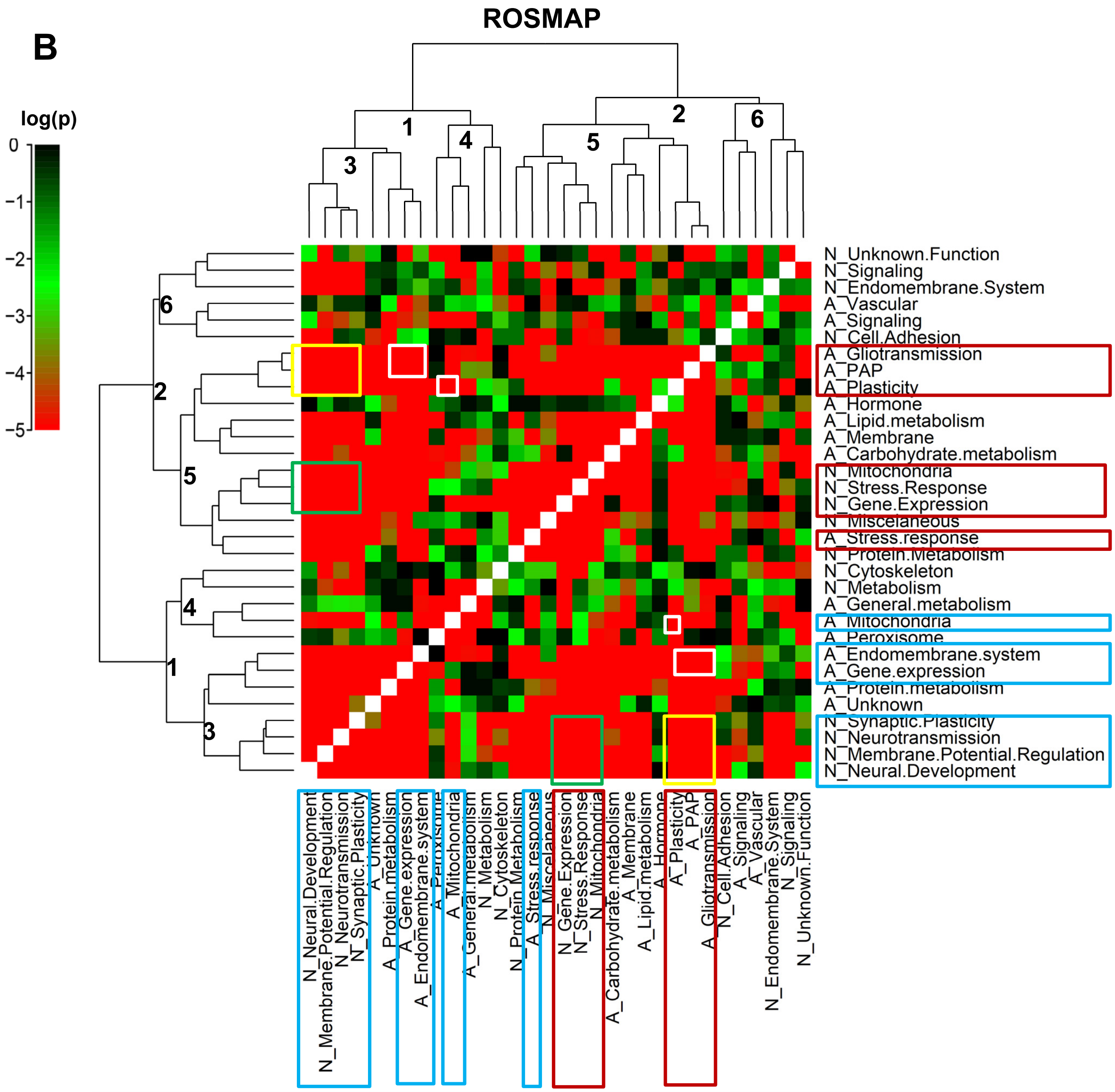

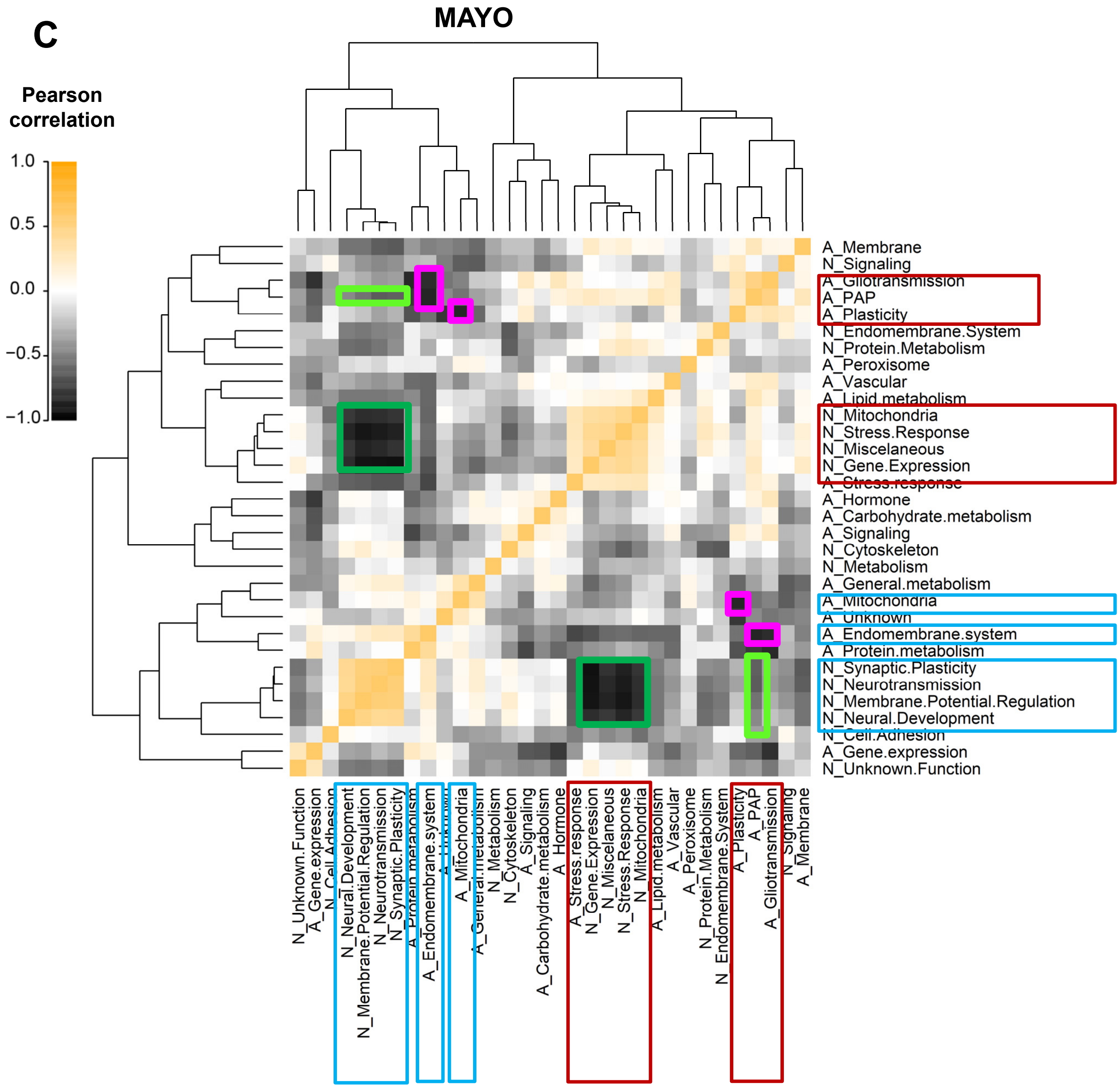

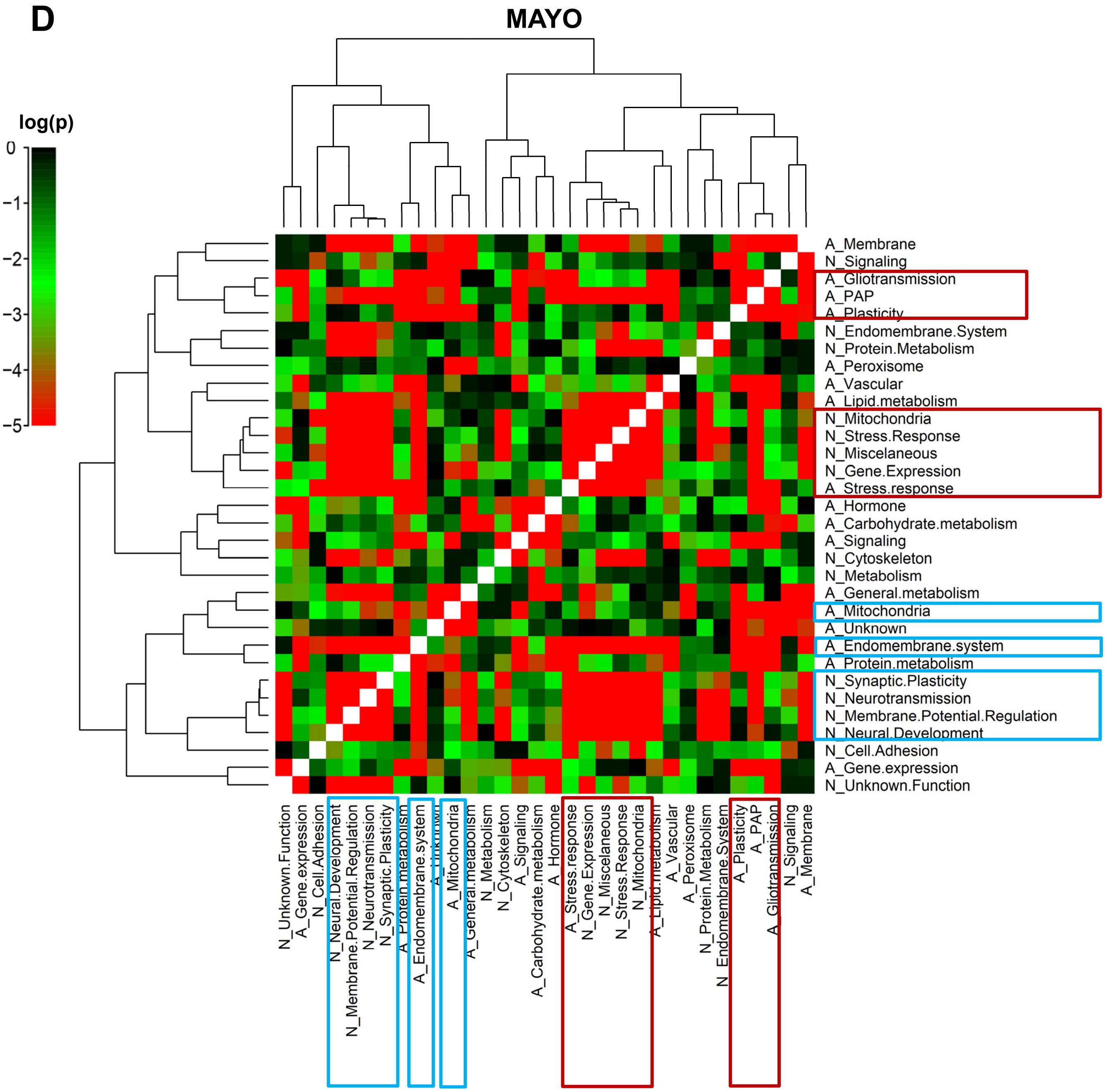

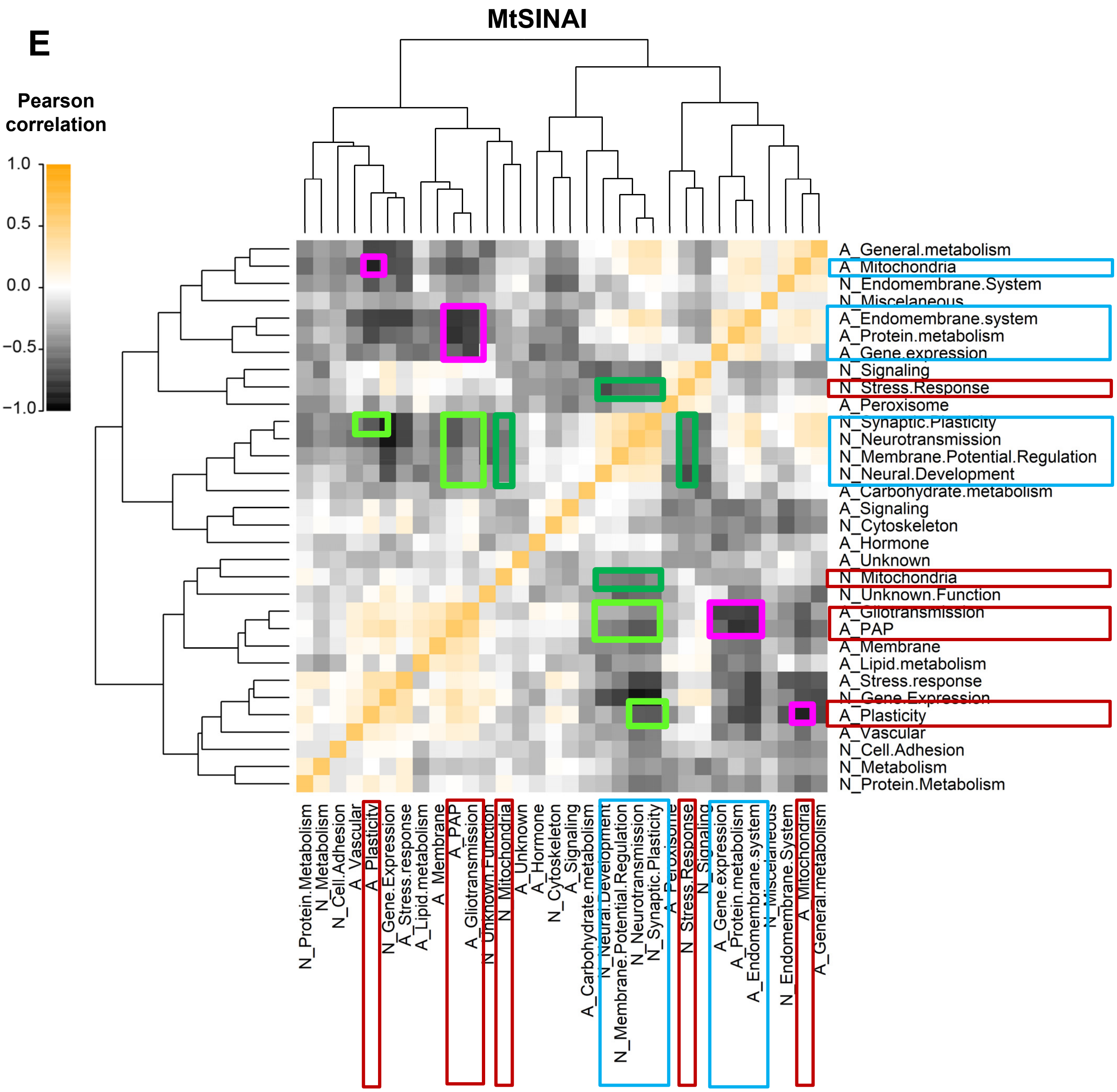

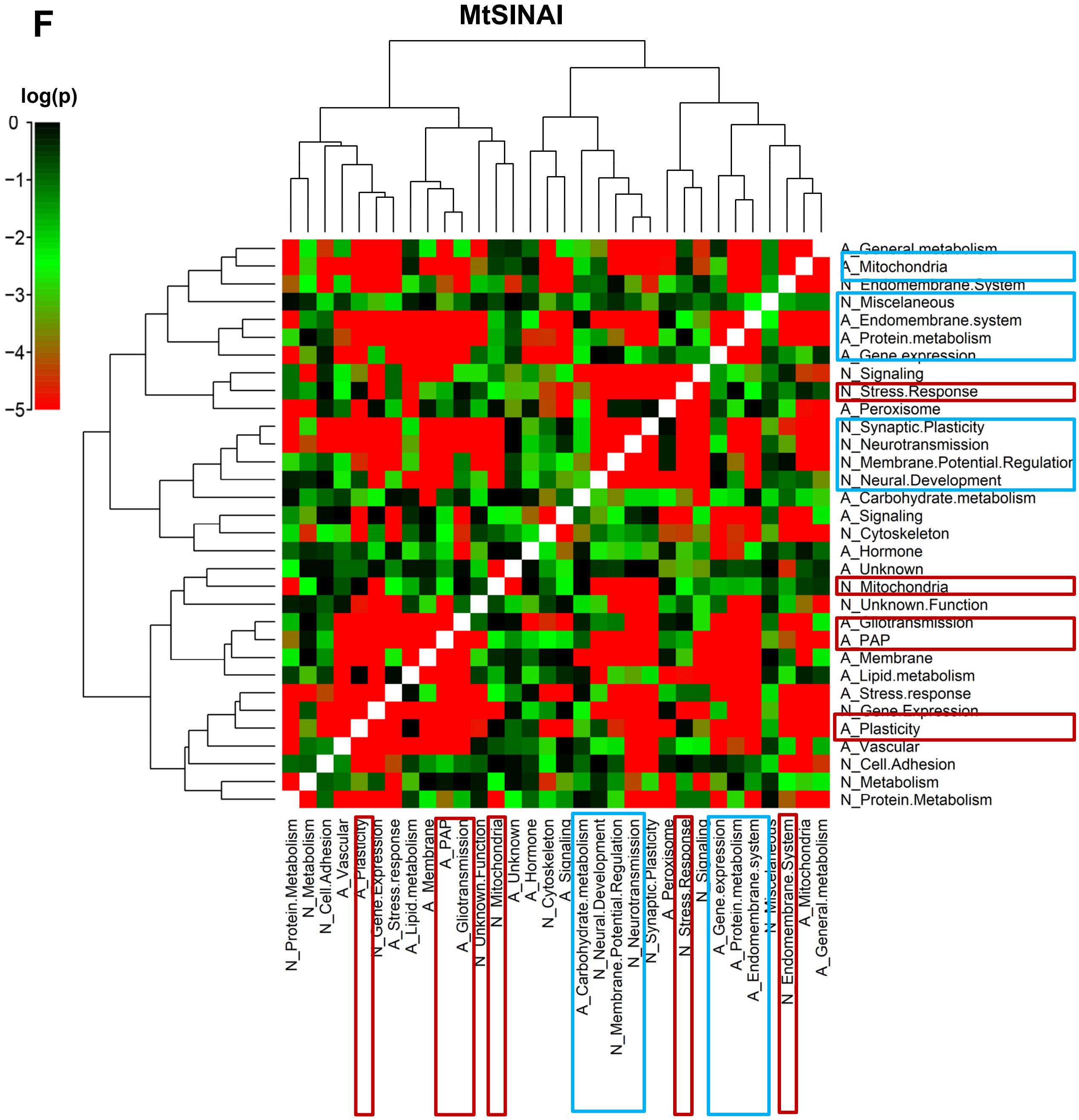
